# The long non-coding RNA *TRIB1AL* links metabolic dysfunction-associated steatotic liver disease, cardiometabolic risk and human lifespan

**DOI:** 10.1101/2025.06.11.25329423

**Authors:** Émilie Gobeil, Jérôme Bourgault, Eloi Gagnon, Nolwenn Samson, Louis-Jacques Ruel, Valérie Côté, Patricia L. Mitchell, Christian Couture, Clarisse Gotti, Florence Roux-Dalvai, Arnaud Droit, Yohan Bossé, Patrick Mathieu, Fannie Lajeunesse-Trempe, Marie-Claude Vohl, Alexandre Caron, André Tchernof, Sébastien Thériault, Mathieu Laplante, Benoit J. Arsenault

**Affiliations:** Centre de recherche de l’Institut universitaire de cardiologie et de pneumologie de Québec - Université Laval, Québec (QC), Canada; Proteomics Platform, CHU de Québec - Université Laval Research Center, Québec (QC), Canada; Department of Molecular Medicine, Faculty of Medicine, Université Laval, Québec (QC), Canada; Department of Surgery, Faculty of Medicine, Université Laval, Québec (QC), Canada; Faculté de pharmacie, Université Laval, Québec (QC), Canada; School of Nutrition, Université Laval, Québec (QC), Canada; Centre Nutrition, santé et société, Institut sur la nutrition et les aliments fonctionnels, Université Laval, Québec (QC), Canada; Department of Molecular Biology, Medical Biochemistry and Pathology, Faculty of Medicine, Université Laval, Québec (QC), Canada; Department of Medicine, Faculty of Medicine, Université Laval, Québec (QC), Canada

## Abstract

Genome-wide association studies (GWAS) have identified dozens of genetic loci linked with metabolic dysfunction-associated steatotic liver disease (MASLD). To identify liver-expressed genes that may represent therapeutic candidates for MASLD, we conducted a new GWAS meta-analysis including 16,532 cases and 1,240,188 controls, as well as RNA sequencing of liver samples and genome-wide genotyping of 504 individuals of the Quebec Obesity Biobank. Using Mendelian randomization (MR) and genetic colocalization, we confirm the implication of genes previously linked with MASLD and identified novel ones including *AKNA* (AT-hook transcription factor), *EPHA2* (EPH receptor A2), *CHEK2* (encoding Checkpoint kinase 2) and *PCCB* (Propionyl-CoA carboxylase subunit beta). More specifically, we found a strong and positive effect of the long non-coding RNA *TRIB1AL* on MASLD. The lead genetic variant was not linked with expression levels of the nearby protein-coding gene *TRIB1* (Tribbles Pseudokinase 1). In participants of the UK Biobank with whole exome sequencing data available, rare loss-of-function variants in *TRIB1* were not associated with liver fat accumulation or plasma triglyceride levels, suggesting that the long non-coding RNA *TRIB1AL* may carry cardiometabolic effects independently of *TRIB1*. Targeted- and phenome-wide MR also identified lower liver-expressed *TRIB1AL* as being associated with reduced liver fat accumulation, lower plasma lipoprotein-lipid levels, decreased atherosclerotic cardiovascular disease risk, and increased human lifespan. These results open the door to liver-targeted therapeutics silencing of the non-coding genome for the prevention and treatment of MASLD and cardiometabolic diseases.

---

Metabolic dysfunction-associated steatotic liver disease (MASLD, formerly known as non-alcoholic fatty liver disease [NAFLD]) is now affecting up to one in three adults and one in ten children worldwide.^1–4^ Understanding the pathogenesis of MASLD and identifying preventive strategies for this disease and its associated complications are urgent public health priorities.

Large-scale genome-wide association studies (GWAS) by our group and others^5–9^ have identified dozens of genetic loci linked to MASLD, including *PNPLA3* (Patatin like phospholipase domain containing 3), *TM6SF2* (Transmembrane 6 superfamily member 2), and more recently, *TRIB1* (Tribbles pseudokinase 1), shedding light on the genetic architecture of this disease and identifying potential therapeutic targets. While GWAS identify genetic variants associated with traits, they do not necessarily pinpoint the causal genes. Moreover, most significant GWAS signals are located in non-coding regions with suspected regulatory functions that are challenging to interpret.^10^

The leading cause of mortality observed in patients with MASLD is atherosclerotic cardiovascular diseases (ASCVD), suggesting that therapies targeting both liver fat accumulation/MASLD and ASCVD may significantly improve outcomes and quality of life for patients.^11^ Understanding the genetic and molecular determinants of MASLD is essential for developing effective strategies to mitigate its burden, ultimately reducing the risk of liver-related complications and ASCVD in affected individuals.

Here, we generated a new dataset of liver expression quantitative trait loci (eQTL) and performed a large GWAS meta-analysis of electronic health record-documented MASLD in participants of European ancestry to identify liver expressed genes that may cause MASLD and its related complications. Using a comprehensive Mendelian randomization (MR) strategy, we identified the long non-coding RNA *TRIB1AL* (*TRIB1* associated lncRNA) as a significant predictor of MASLD. The objective was to characterize the health effects of *TRIB1AL* and to evaluate the potential of this long non-coding RNA as a therapeutic target for MASLD and cardiometabolic diseases.

## Results

### Identification of liver-expressed genes associated with MASLD

A recent GWAS performed in the Million Veteran Program identified 98 genetic loci associated with MASLD using a proxy MASLD definition of chronic elevation of alanine aminotransferase (ALT) levels without other liver diseases.^7^ In order to identify liver-expressed genes that may potentially be causally associated with MASLD, we performed a combination of MR and genetic colocalization using as study exposures genes located within these 98 loci, and MASLD as the main study outcome. To derive our study exposures, we performed RNA sequencing of liver samples obtained from 504 patients undergoing bariatric surgery (Quebec Obesity Biobank) and mapped expression quantitative trait loci (eQTLs), as previously described.^12^ The characteristics of the participants of the Quebec Obesity Biobank are presented in Supplementary Table 1.To derive the study outcome, a new GWAS meta-analysis of seven cohorts totaling 16,532 cases and 1,240,188 controls was performed (Supplementary Table 2). The Manhattan plot of this genome-wide meta-analysis is presented in Supplementary Figure 1. Figure 1 and Supplementary Table 3 present the effect of genetically predicted liver-expressed genes at MASLD loci on MASLD. This analysis identified several genes previously associated with MASLD such as *PNPLA3*, *TM6SF2*, *MTTP* (encoding Microsomal triglyceride transfer protein), *GPAM* (encoding Glycerol-3-phosphate acyltransferase 1), *PPP1R3B* (encoding Protein phosphatase 1 regulatory subunit 3B), *APOH* (encoding Apolipoprotein H) and *COBLL1* (encoding Cordon-bleu protein-like 1). Genes not previously reported to be associated with MASLD such as *AKNA* (encoding AT-hook transcription factor), *EPHA2* (encoding EPH receptor A2), *CHEK2* (encoding Checkpoint kinase 2), *PCCB* (encoding Propionyl-CoA carboxylase subunit beta) were also identified. This analysis also revealed that the long non-coding RNA *TRIB1AL* had the most significant effect on MASLD with higher expression of *TRIB1AL* being positively associated with MASLD.

**Figure 1.**
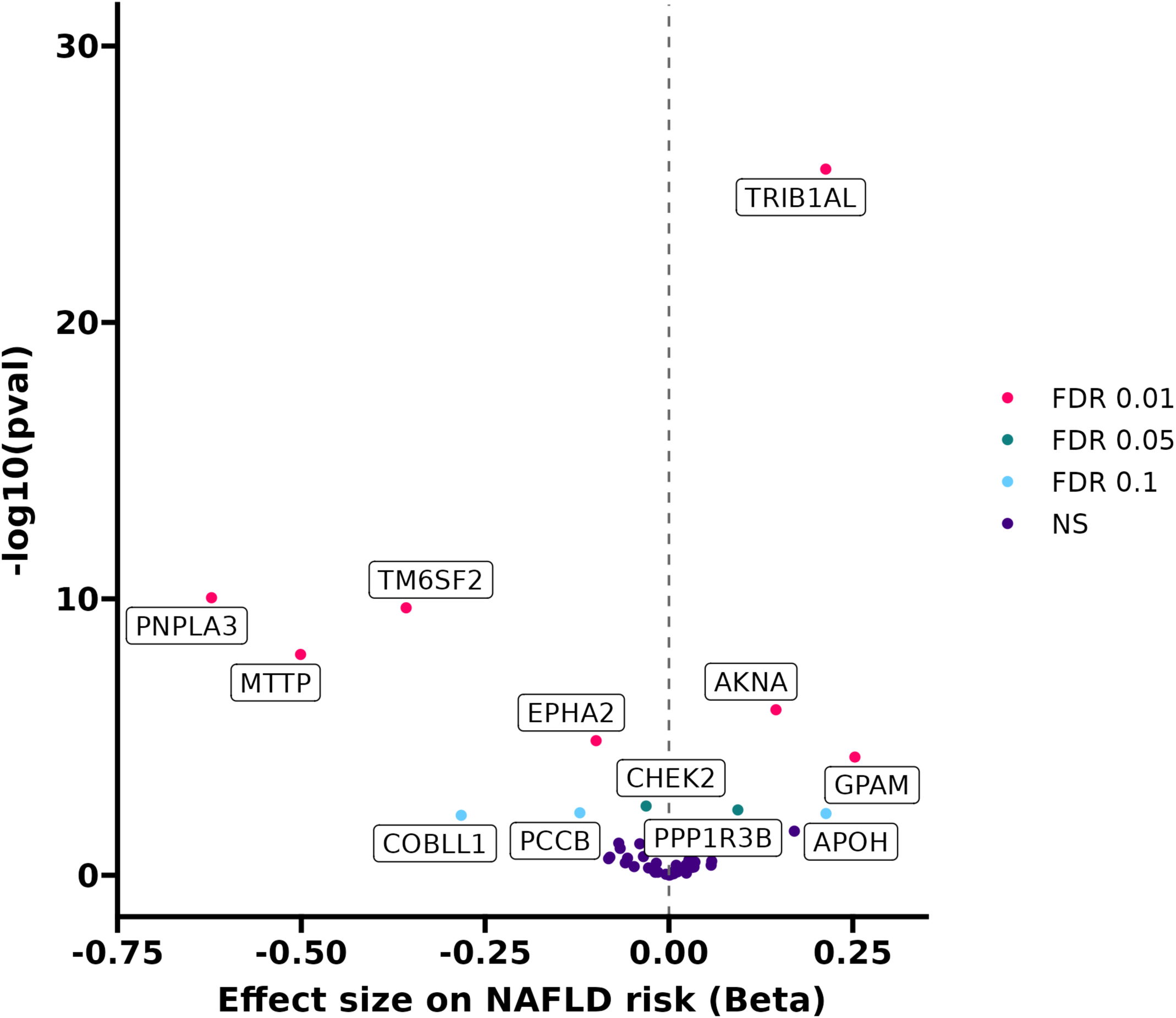
Volcano plot presenting the impact of genetically predicted liver gene expression levels at know metabolic dysfunction-associated steatotic liver disease (MASLD) genome-wide association study loci. Two-sample mendelian randomization analyses between genes located within known MASLD loci as study exposure and MASLD as the study outcome. Each liver-expressed gene with at least one eQTL genome-wide significant was used as exposure. A meta-analysis of GWAS totaling 16,532 cases and 1,240,188 controls was used for the study outcome.

### Fine-mapping at the TRIB1/TRIB1AL locus

Previous GWAS have identified a signal at the *TRIB1/TRIB1AL* locus for plasma lipids, MASLD and coronary artery disease (CAD)^5,13,14^ but the gene responsible for association is still debated. Although our initial finding revealed a potentially causal effect of *TRIB1AL*, we further investigated the respective contribution of *TRIB1AL* and *TRIB1* to MASLD. Supplementary Figure 2 presents the locuszoom plots revealing that the MASLD signal may lie downstream of *TRIB1* within the *TRIB1AL* locus. Conditional analyses performed using the Pair-Wise Conditional analysis and Colocalisation (PWCoCo) analysis also revealed that *TRIB1AL* remained associated with MASLD upon conditioning for *TRIB1*, with a posterior probability H4 (PPH4) of 0.993. The lead genetic variant identified by PWCoCo is rs28601761. This variant was dose-dependently associated with liver expression of *TRIB1AL* but not *TRIB1* (Figure 2) in the Quebec Obesity Biobank. This variant was also associated with liver expression of *TRIB1AL* in the STARNET browser^15^ but not in the Genotype-Tissue Expression (GTEx) project^16,17^ (data not shown). The effect of this variant on *TRIB1AL* expression was consistent across MASLD stages (Supplementary Figure 3). To assess the potential tissue specificity of *TRIB1AL*, we reckoned the tissue-specific gene expression metric Tau. Results presented in Supplementary Figure 4 revealed higher expression levels of *TRIB1AL* in the liver compared to other tissues (Tau = 0.94). *TRIB1AL* was also expressed at lower levels in the kidney cortex and the thyroid.

**Figure 2.**
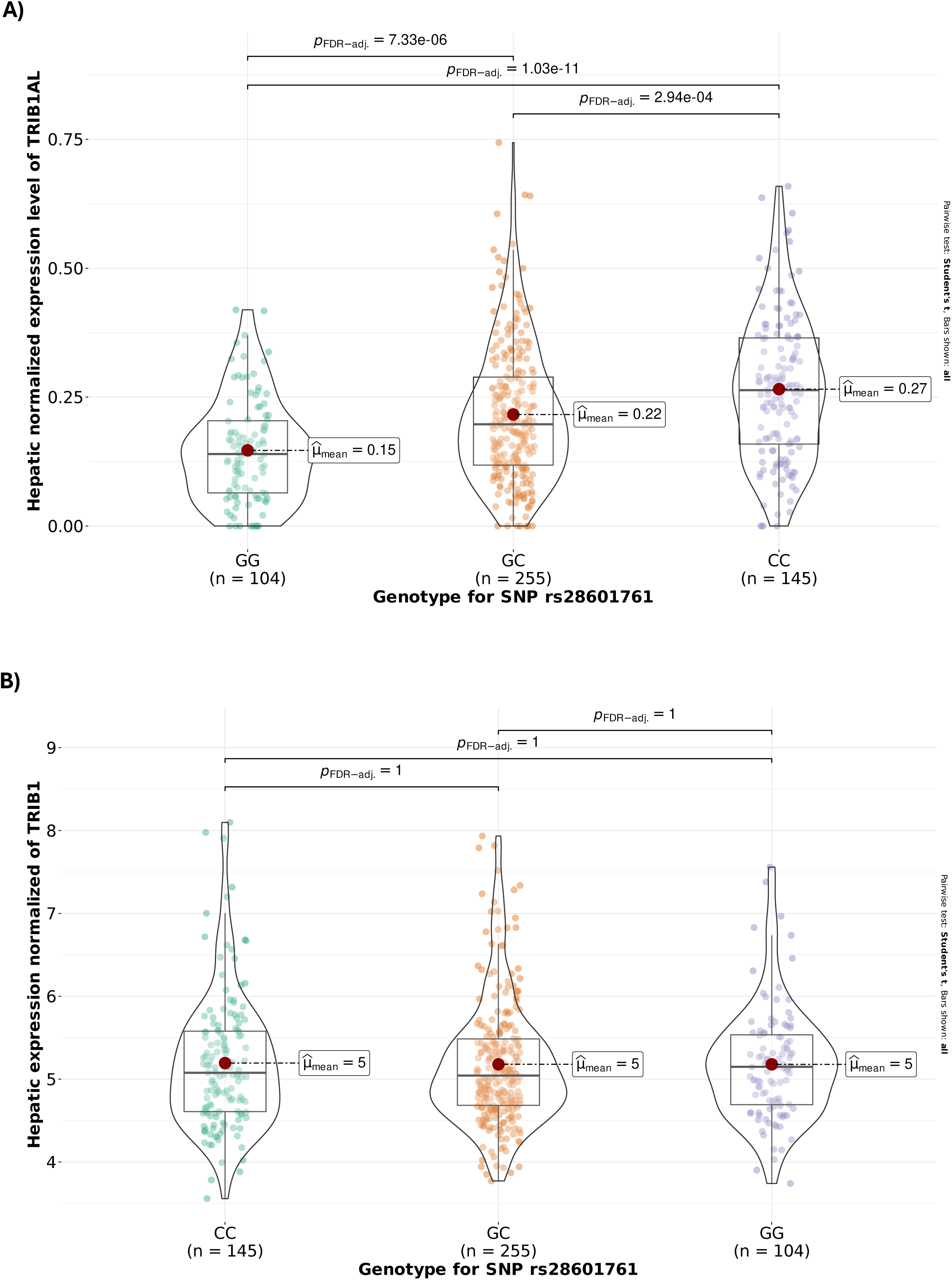
Impact of the causal variant at the *TRIB1* locus on liver *TRIB1* and *TRIB1AL* expression in participants of the Quebec Obesity Biobank. rs28601761 was associated with A) *TRIB1AL,* but not with B) *TRIB1* liver expression levels. A total of 504 participants of the Quebec Obesity Biobank were separated according to rs28601761 genotype. Normalized gene expression levels of liver *TRIB1* and *TRIB1AL* are presented as log2(TPM+1) and were compared using Student t-tests.

### Cardiometabolic effects of protein-truncating variants at TRIB1

Given that *TRIB1* is a protein-coding gene, we aimed to investigate whether *TRIB1* was associated with cardiometabolic traits by assessing the impact of protein-truncating variants (PTVs) in *TRIB1* on plasma triglyceride levels and liver fat accumulation in participants of the UK Biobank. We used PTVs at *TM6SF2*, another protein-coding gene known to be functionnaly associated with these traits^18,19^, as a positive control. Results presented in Figure 3 show that, unlike PTVs in *TM6SF2*, PTVs in *TRIB1* are not associated with plasma triglycerides or liver fat accumulation. Taken together, these results show that the protein-coding gene *TRIB1* may not be functionnaly linked with cardiometabolic traits.

**Figure 3.**
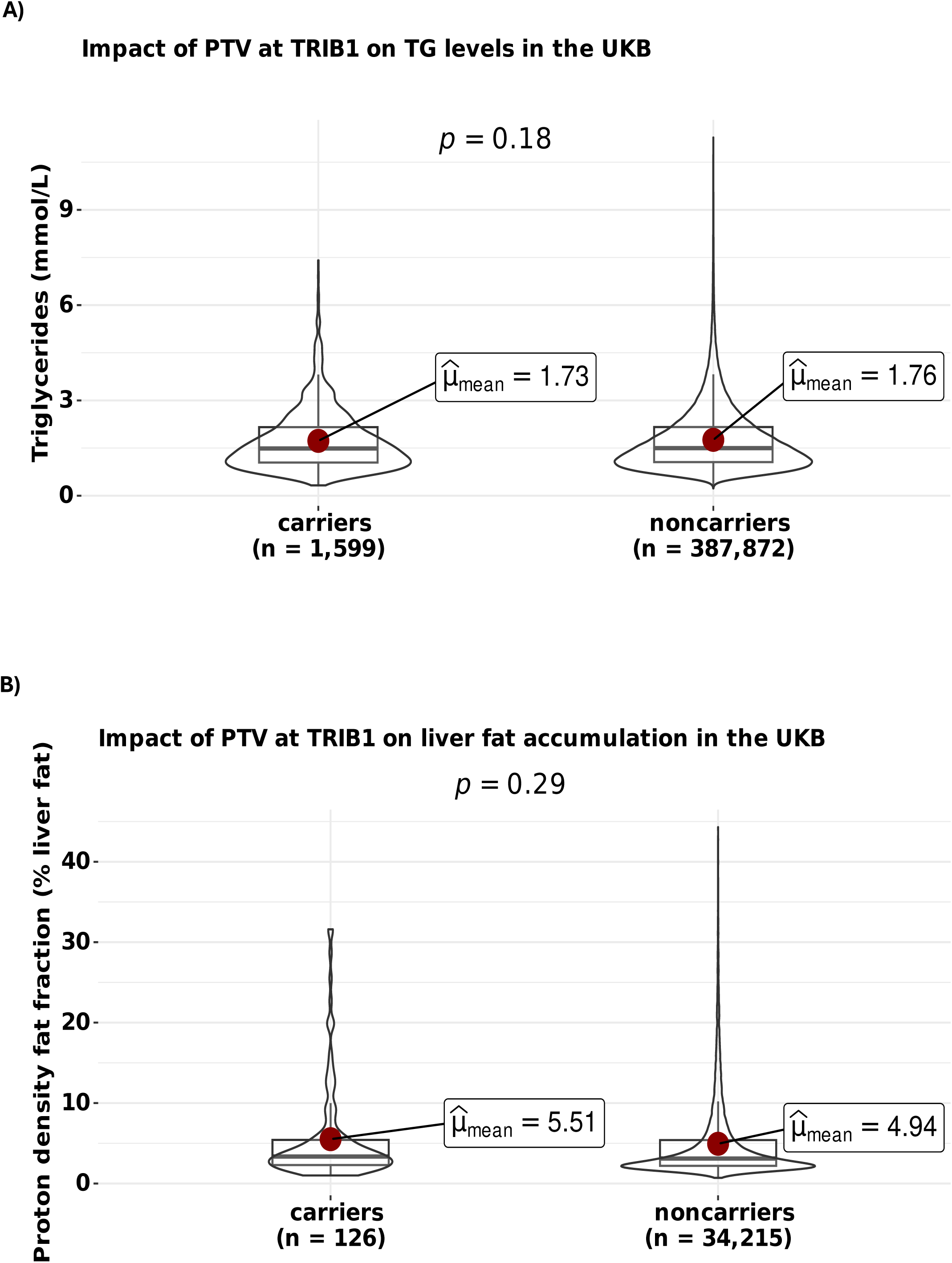
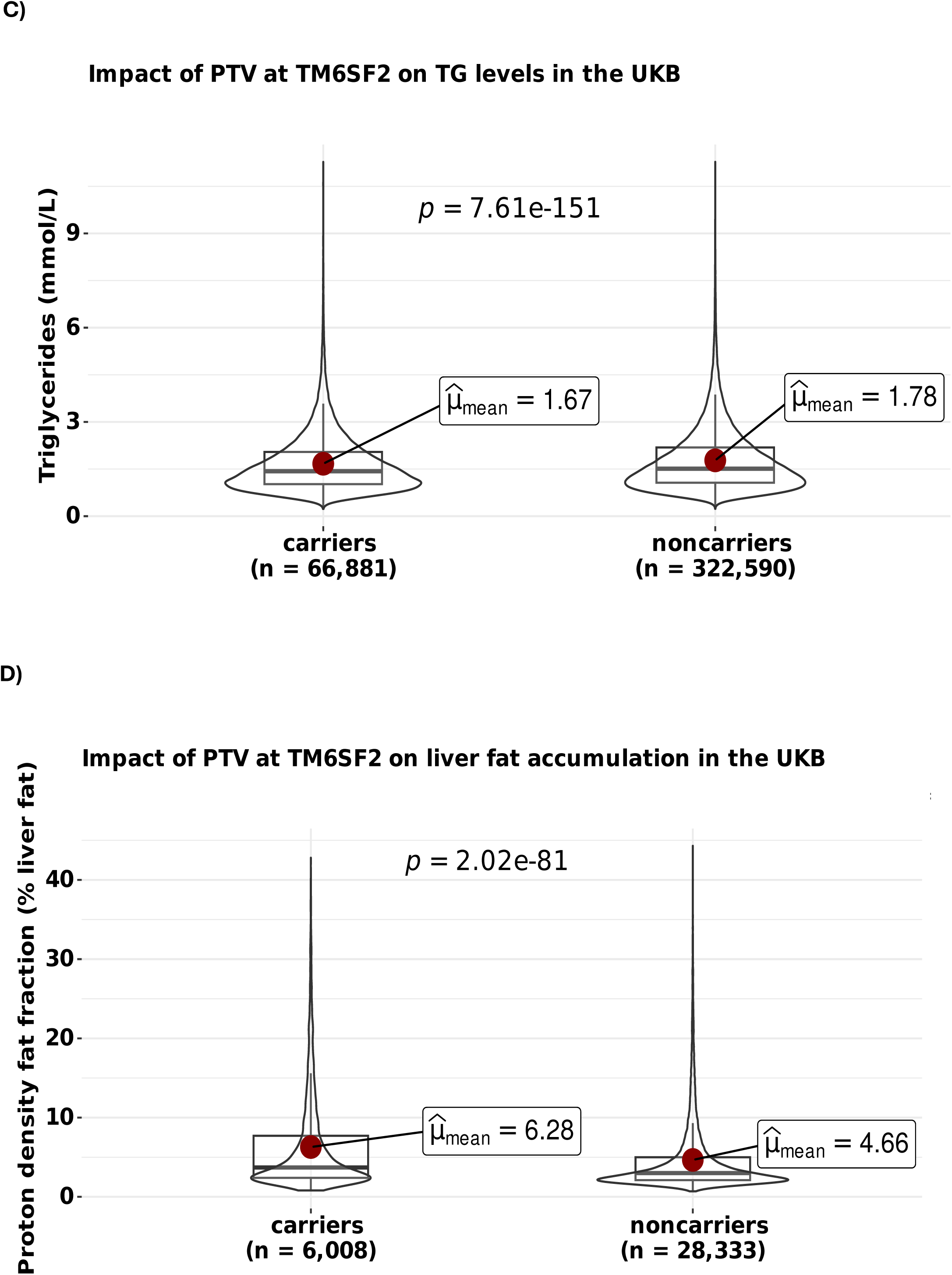
Impact of *TRIB1* and *TM6SF2* protein-truncating variants on triglyceride levels and liver fat accumulation in participants of the UK Biobank. Carriers of at least one *TRIB1* protein-truncating variant (PTV) had comparable triglyceride levels (A) and liver fat accumulation (B) compared to noncarriers. Carriers of at least one *TM6SF2* protein-truncating variant (PTV) had higher triglyceride levels (C) and higher liver fat accumulation (D) compared to noncarriers. Triglyceride and liver fat accumulation levels between carriers and noncarriers were compared using Student t-tests.

### Health effects of genetically predicted reductions in hepatic TRIB1AL expression

We used MR to predict the impact of genetically predicted liver *TRIB1AL* “inhibition” on a wide range of cardiometabolic traits and diseases (Supplementary Table 4) to determine if *TRIB1AL* may represent a new therapeutic target for MASLD and cardiometabolic diseases. We compared the effects of *TRIB1AL* to *ANGPTL3* (Angiopoietin like 3) and *PCSK9* (Proprotein convertase subtilisin/kexin type 9), two liver-specific drug targets under investigation for cardiometabolic diseases. A one-standard decrease in genetically predicted liver *TRIB1AL* expression (mimicking *TRIB1AL* inhibition) was associated with a more favorable cardiometabolic risk profile (Figure 4A). Compared to genetic inhibition of *ANGPTL3* and *PCSK9*, a 1-SD lowering in *TRIB1AL* was more strongly associated liver fat accumulation, alanine aminotransferase (ALT) and high-density lipoprotein (HDL) cholesterol levels. Genetic inhibition of *TRIB1AL* was associated with lower apolipoprotein B levels, similar to genetic inhibition of *ANGPTL3* and *PCSK9*, although the latter showed slightly more potent effects. Genetic inhibition of *TRIB1AL* had beneficial effects on triglyceride levels, albeit to a lower extent than genetic inhibition of *ANGPTL3*.

**Figure 4.**
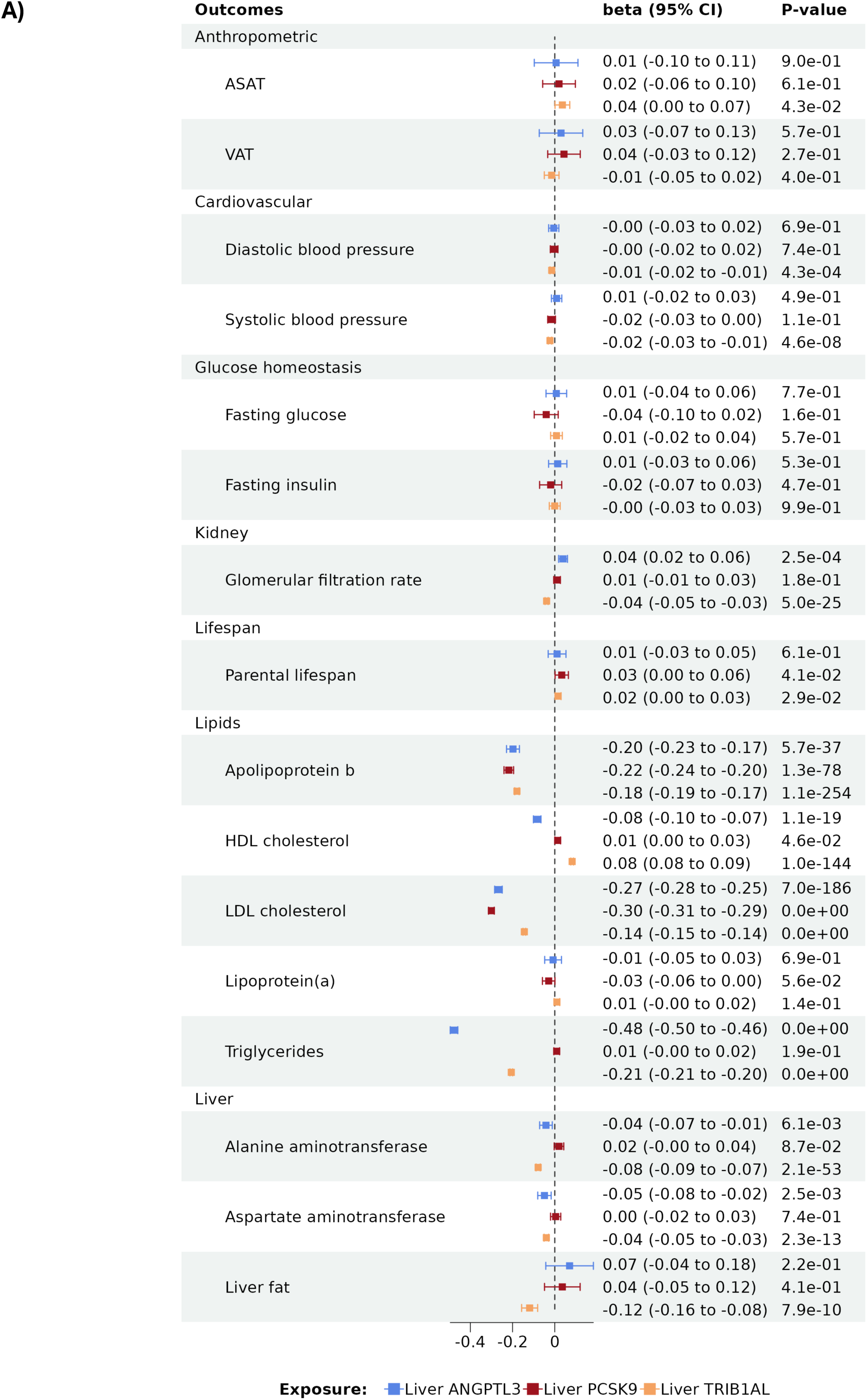
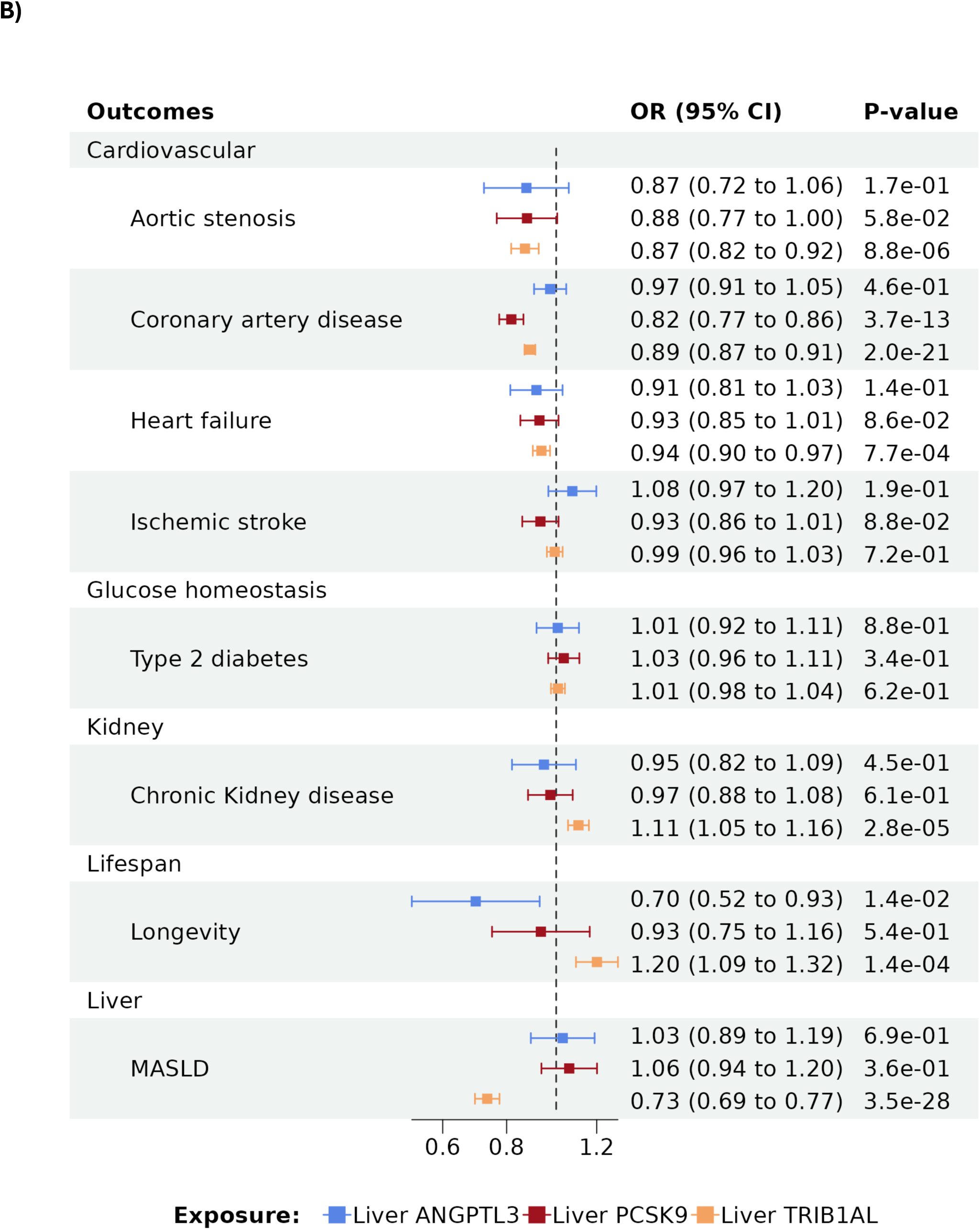
Impact of genetically predicted reductions in liver expression of *TRIB1AL*, *ANGPTL3* and *PCSK9* on cardiometabolic phenotypes and diseases. Effects of the strongest SNP associated with the liver *TRIB1AL*, *ANGPTL3* and *PCSK9* expression on cardiometabolic A) quantitative outcomes and B) diseases (binary outcomes) was obtained using the Wald ratio. Effect sizes (95% CI) are represented by A) SD or B) log(OR) change in the outcome per 1-SD increase in liver *TRIB1AL*, *ANGPTL3* or *PCSK9* expression. GWAS summary statistics from the largest genome-wide association studies (GWAS) were included as study outcomes.

Genetic inhibition of *TRIB1AL* was also associated with lower systolic and diastolic blood pressure and with lower estimated glomerular filtration rate (eGFR). Genetic inhibition of *TRIB1AL* was also associated with higher parental lifespan. Figure 4B presents the effects of genetically predicted reductions in liver *TRIB1AL*, *ANGPTL3* and *PCSK9* inhibition on cardiometabolic diseases. As expected, genetically predicted reductions in liver *TRIB1AL* were strongly associated with lower MASLD. It was also negatively associated with lower ASCVD (aortic stenosis, CAD and heart failure). However, the impact of genetically predicted reduction in liver *PCSK9* on CAD seemed to be more potent. Consistent with the effect on parental lifespan, genetically predicted reduction in liver *TRIB1AL* was also associated with increased longevity. Also, consistent with the effect of genetically predicted reduction in liver *TRIB1AL* on lower eGFR and possibly the expression of *TRIB1AL* in kidney cortex, genetically predicted reduction in liver *TRIB1AL* seemed to be linked with higher chronic kidney disease (CKD).

Given the overall positive effects of genetically predicted reductions in liver *TRIB1AL* on MASLD, cardiometabolic risk and lifespan, liver *TRIB1AL* inhibition may represent a therapeutic target of interest for cardiometabolic diseases.

### Phenome-wide association study of hepatic TRIB1AL expression

To better understand the health effects of *TRIB1AL* and to document potential health benefits or hazards associated with liver *TRIB1AL* inhibition, we performed a phenome-wide MR study. Supplementary Figure 5 presents the health effect of the previously identified lead genetic variant within *TRIB1AL* (rs28601761) on 330 human diseases or phenotypes in the pooled analysis of FinnGen, UK Biobank and the Million Veteran Program. Results of this investigation confirmed the effect of genetically predicted reductions in liver *TRIB1AL* expression on MASLD-related diseases and revealed potential reductions in gastrointestinal tract-related cancers. As expected, this analysis showed that genetically predicted reduction in liver *TRIB1AL* expression was associated with a lower risk of hypercholesterolemia and ASCVD-related outcomes. Genetically predicted reduction in liver *TRIB1AL* expression was however associated with higher risk of cholelithiasis (gallstones) and cholecystitis. Altogether, the assessment of genetically predicted reductions in liver *TRIB1AL* expression revealed mostly beneficial effects on cardiometabolic traits and diseases.

### Assessing TRIBAL effects through multiomic Mendelian randomization

Genes and proteins influenced by *TRIB1AL* may identify direct or indirect targets of this lncRNA and shed light on its mechanisms of action. To gain knowledge on the liver and systemic effects of lower liver *TRIB1AL* expression, we performed additional MR analyses using the lead SNP as study exposure and the Quebec Obesity Biobank transcriptome as study outcome (Methods). These analyses revealed that *TRIB1AL* expression was negatively associated with known genes influencing MASLD such as *TM6SF2* and *TRHDE (*Thyrotropin releasing hormone degrading enzyme) (Figure 5A). Analyses of homozygote carriers of the risk and protective allele (CC versus GG carriers of rs28601761) did not reveal additional effects of *TRIB1AL* on the liver transcriptome (data not shown).

**Figure 5.**
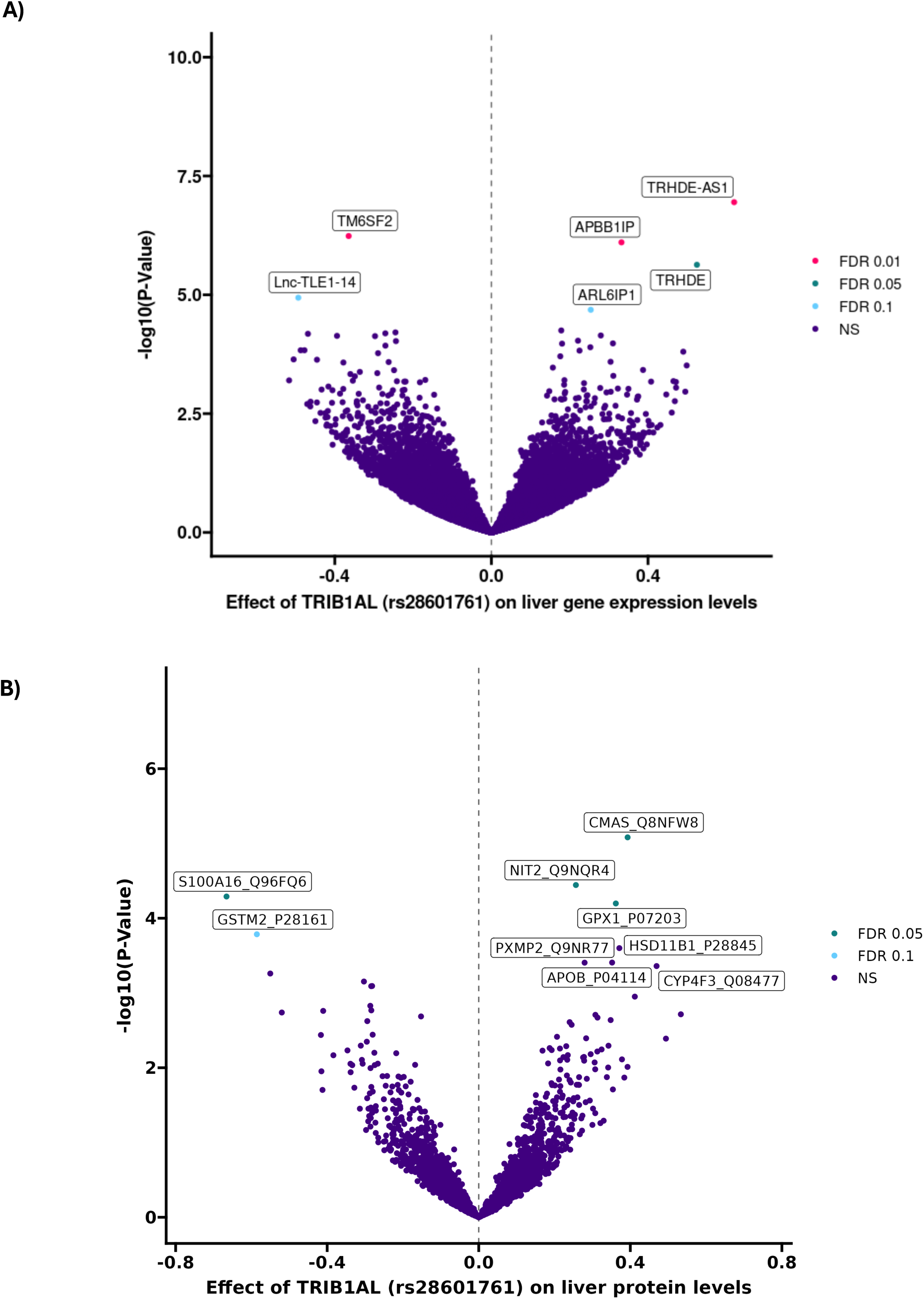
Impact of genetically predicted liver *TRIB1AL* expression on the liver transcriptome and proteome. Effects of the strongest SNP associated with the liver *TRIB1AL* expression on the liver transcriptome (A) and the liver proteome (B) in patients of the Quebec Obesity Biobank.

Assessing protein concentrations in addition to gene expression levels is critical, especially in light of recent studies describing a rather poor correlation between gene expression and protein levels.^20,21^ Because proteins are usually the primary effector of biological function, their inclusion in post-GWAS analyses may improve our understanding of the biological mechanisms influencing MASLD. We therefore measured approximately 3,000 liver proteins using LC-MS/MS and performed additional MR analyses using our new liver proteomic dataset (n = 460). These analyses revealed that genetically predicted liver *TRIB1AL* expression was associated with proteins that may be linked with MASLD such as apolipoprotein B (*APOB*), hydroxysteroid 11-beta dehydrogenase 1 (*HSD11B1*), S100 calcium binding protein A16 (S100A16), glutathione S-transferase Mu2 (*GSTM2*) and glutathion peroxidase 1 (GPX1) (Figure 5B). Analyses of homozygote carriers of the risk and protective allele (CC versus GG carriers of rs28601761) did not reveal additional effects of *TRIB1AL* on the liver proteome (data not shown).

The impact of genetically predicted liver *TRIB1AL* expression was also tested on over 1,500 blood proteins measured by Olink proximity extension assay in >50,000 participants of the UK Biobank. Several blood proteins were positively associated genetically predicted liver *TRIB1AL* higher expression such as Interleukin 1 Receptor Type 2 (IL1R2) and Fetuin B (FETUB) (Figure 6A). Gene ontology analyses from these proteins were performed to identify biological pathways that may be altered by liver *TRIB1AL* expression (Figure 6B). All identified pathways related to lipoprotein-lipid metabolism. Some terms were also related to blood coagulation. Altogether, these new results provide insight into the role of *TRIB1AL* on liver metabolism and identified potential genes, proteins and pathways that may be directly or indirectly influenced by *TRIB1AL*.

**Figure 6.**
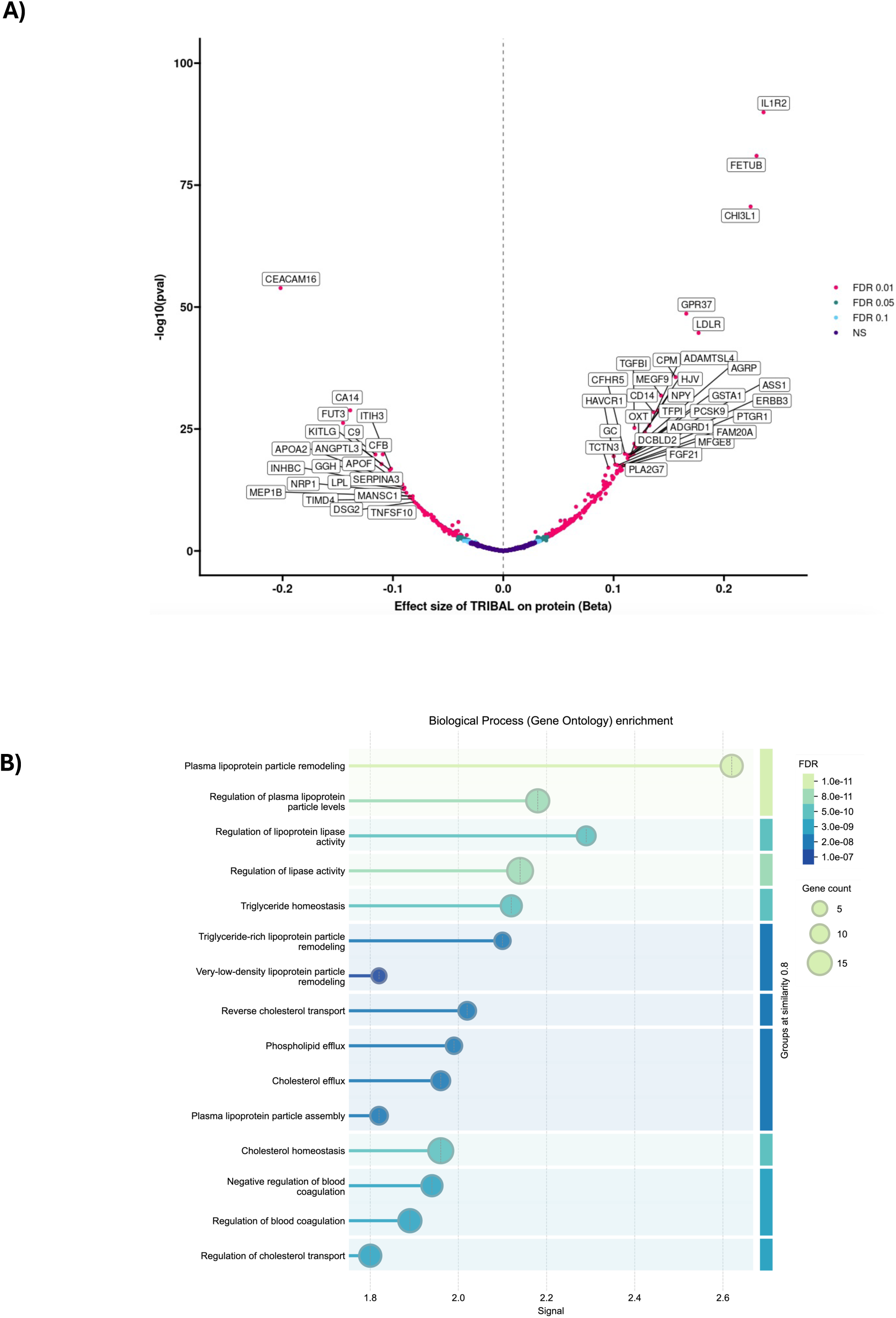
Impact of genetically predicted liver *TRIB1AL* expression on the blood proteome. A) Effects of the strongest SNP associated with the liver *TRIB1AL* expression on blood proteins was obtained using the Wald ratio. Effect sizes (95% CI) are represented by SD change in the outcome per 1-SD increase in liver TRIB1AL expression. GWAS on over 1500 blood proteins measured by Olink proximity extension assay in >50,000 participants of the UK Biobank were used as study outcomes. B) Biological process (Gene ontology) enrichment of blood protein impacted significantly by the strongest SNP associated with the liver *TRIB1AL* expression.

## Discussion

Through integrative genomics leveraging a new GWAS meta-analysis and liver eQTLs and pQTLs from approximately 500 participants from the Quebec Obesity Biobank, we identified novel genes linked with MASLD and demonstrate that *TRIB1AL*, rather than the adjacent protein-coding gene *TRIB1*, is a likely causal effector at this locus. Using MR, genetic colocalization, and phenome-wide association analyses, we provide converging evidence that genetic inhibition of liver *TRIB1AL* is associated with lower liver fat accumulation, improved lipoprotein-lipid profiles, reduced cardiometabolic disease risk, and potentially increased human lifespan.

These findings challenge long-standing assumptions about *TRIB1* being the principal effector gene at this locus.^22^ In one of the earliest GWAS of lipoprotein-lipid levels, Kathiresan et al.^23^ identified a signal at the *TRIB1* locus associated with triglyceride levels. Functional analyses at this locus later revealed a complex interplay between *TRIB1* and *TRIB1AL* in the regulation of triglyceride levels.^24^ Although *TRIB1* has been widely implicated in lipid metabolism and cardiovascular risk based on GWAS hits, our analyses of PTVs in the UK Biobank reveal no associations between *TRIB1* loss-of-function and liver fat or plasma triglycerides. In contrast, genetically predicted higher *TRIB1AL* expression is consistently linked to adverse cardiometabolic outcomes, positioning *TRIB1AL* as a compelling new therapeutic target.

Importantly, we show that *TRIB1AL* is predominantly expressed in the liver and displays tissue specificity consistent with a hepatic regulatory function. Conditional colocalization analyses further support that the genetic variant rs28601761, associated with MASLD and lipids, regulates *TRIB1AL* but not *TRIB1* expression. Although these findings need to be further validated with functional experiments, the present results suggest that *TRIB1AL* may function independently of *TRIB1*, representing a distinct layer of post-GWAS biology involving the non-coding genome. These results are consistent with the recent work of Soubeyrand et al.^25^

In addition to its role in MASLD, *TRIB1AL* inhibition appears to have a broader beneficial cardiometabolic profile, comparable in magnitude, and in some cases, superior to inhibition of established drug targets such as *PCSK9* and *ANGPTL3*. Specifically, genetically predicted *TRIB1AL* inhibition was associated with reductions in plasma apolipoprotein B and triglycerides, blood pressure, and risk of ASCVD, while improving lifespan. These effects underscore the potential for *TRIB1AL*-targeted therapies to simultaneously address multiple facets of metabolic disease. The phenome-wide MR further highlights both the promise and complexity of targeting *TRIB1AL*. While reductions in its expression correlate with lower risk of MASLD, cardiovascular disease, and certain gastrointestinal cancers, they are also associated with a modestly increased risk of cholelithiasis and chronic kidney disease (although the later may be explained by local effect of *TRIB1AL* in the kidney). Whether liver specific inhibition of *TRIB1AL* could influence kidney function is unknown. Regardless, this potential trade-off will need to be considered in therapeutic development.

To elucidate the mechanisms through which *TRIB1AL* influences hepatic and systemic physiology, we performed multiomic MR using transcriptomic and proteomic datasets. Although functional studies will need to be performed to assess the main target(s) of *TRIB1AL*, our transcriptomic and proteomic analyses revealed that *TRIB1AL* may regulate expression levels of many genes and proteins involved in liver fat accumulation and lipoprotein metabolism such as *TM6SF2* and apolipoprotein B. Proteomic MR in blood samples revealed that potential downstream targets include PCSK9, LDLR and LPL. The circulating proteins most strongly associated with *TRIB1AL* were IL1R2 and Fetuin B (FETUB), which are implicated in inflammation and lipid metabolism. Interestingly, antisense oligonucleotide-mediated inhibition of *TRIB1AL* was previously shown to reduced Fetuin B.^25^ Gene ontology analyses of these proteins further suggest that *TRIB1AL* may exert its effects through pathways related to lipoprotein-lipid homeostasis, though the precise mechanisms, whether direct or mediated through other RNA-binding or epigenetic processes, remain to be clarified.

The present study has several strengths, including the use of a large, well-powered GWAS meta-analysis, a new deeply phenotyped liver expression cohort, and comprehensive MR and colocalization pipelines. However, we also recognize limitations. First, while genetic proxies offer strong causal inference, they cannot fully recapitulate pharmacologic inhibition. Second, *TRIB1AL*’s molecular function whether it acts via RNA, protein interactions, chromatin remodeling, or other mechanisms remains unknown and warrants further study. Third, since our analyses focused on European ancestry populations, replication in diverse ancestries will be essential for equitable translation.

In conclusion, the current study identified *TRIB1AL* as a liver-enriched lncRNA with potentially causal effects on MASLD and cardiometabolic health. These findings provide support for a new model in which non-coding RNA transcripts, rather than nearby protein-coding genes, may be the true effectors at GWAS loci. Moving forward, *TRIB1AL* inhibition represents a promising therapeutic avenue for MASLD and related diseases. Future efforts should aim to characterize the molecular functions of *TRIB1AL* and explore RNA-targeting strategies such as liver-targeted antisense oligonucleotides for clinical translation.

## Methods

### Assessment of liver expression quantitative trait loci

Patients undergoing bariatric surgery at the Quebec Heart and Lung Institute (*Institut universitaire de cardiologie et de pneumologie de Québec* [IUCPQ]) provided informed consent to participate to the institutional biobank (Quebec Obesity Biobank). Clinical information at the time of surgery (sex, age, anthropometry, medication use, medical history and comorbidities and glycemic, lipoprotein and liver enzyme profile) was available for all patients. A total of 504 participants passed genotyping and RNA sequencing quality controls (Supplementary Table 1). Liver samples were obtained by incisional biopsy of left lobe and were not cauterized. The grading and staging of histological lesions have been carried out according to the protocol of Brunt et al.^26^ by pathologists who were blind to the objectives of the study. The liver sample procedure and position of the liver sample are standardized for among all surgeons. Total RNA was extracted from 10 mg of frozen liver samples following the manufacturer’s instructions (Qiagen RNeasy Plus). Briefly, tissue was homogenized in 800 µL Qiazol using a VWR Beadmill. A genomic DNA cleaning step was carried out followed by extraction. The concentration and purity (A_260_/A_280_ and A_260_/A_230_) of the samples were determined on a BioTech BioDrop. The quality of the RNA was determined either by BioAnalyzer or estimated by RNA agarose gel. Total RNA was quantified, and quality assessed using a LabChip GXII (PerkinElmer). Libraries were generated from 250 ng of total RNA. The libraries were normalized and pooled and then denatured in 0.05 N NaOH and neutralized using HT1 buffer. The pool was loaded at 225 pM on Illumina NovaSeq S4 lane using Xp protocol as per the manufacturer’s recommendations. The run was performed for 2×100 cycles (paired-end mode). Genomic DNA was extracted from the blood buffy coat using the GenElute Blood Genomic DNA kit (Sigma, St. Louis, MO, USA) and genome-wide genotyping was performed using the GSA-MD array (Illumina, San Diego, CA, USA). Quality controls were done using Illumina Genomestudio software 2.0 with Genotyping module version 2.0.5, with the auto-clustering option and a call rate of 97%. Quality controls were performed using plinkQC. Pre-imputation QC was made with R package plinkQC v0.3.4 and Will Rayner tools. Imputation was performed using the Michigan Imputation Server based on HRC reference panel. Eagle v2.4 and Minimac4 v1.6.3 were used for phasing and imputation respectively. Post-imputation QC steps included the creation of binary plink files with PLINK 2. For RNA sequencing data, alignment to the human reference genome GRCh38/hg38 was performed using STAR v2.5.1, based on the GENCODE v34 annotation. Gene-level expression quantification was based on the GENCODE 34 annotation, collapsed to a single transcript model for each gene using a custom isoform collapsing procedure. Read counts and TPM values were produced with RNA-SeQC v2.4.2.^27^ Gene expression values for all samples were normalized. Covariates for eQTL (expression quantitative trait loci) mapping included sex, age, and the top 10 genotyping principal components. A set of covariates identified using the Probabilistic Estimation of Expression Residuals (PEER) method,^28^ calculated for the normalized expression matrices was also used. For eQTL analyses, expression values were adjusted for 15 PEER factors to improve the number of eGenes discovered. Next, tensorQTL was used for eQTL quantification as described by the developers.^29^ Only SNPs with a minor allele frequency >0.01 were considered for eQTL mapping.

### Genome-wide meta-analysis of MASLD

Most recent publicly available GWAS of non-alcoholic fatty liver disease (NAFLD) were merged through a meta-analysis to obtain a comprehensive set of summary statistics for genetic variants associated with NAFLD. Since the definition of NAFLD was switched to MASLD to better reflect the central role of metabolic dysfunction in fatty liver disease, avoid stigma associated with alcohol, and shift from a diagnosis of exclusion to one of inclusion,^2^ we have decided to use the term MASDL throughout the manuscript. We performed a meta-analysis of GWAS totaling 16,532 cases and 1,240,188 controls from the seven following cohorts (European ancestry): The Electronic Medical Records and Genomics (eMERGE) network, the UK Biobank, the Estonian Biobank, the EPoS Consortium, FinnGen study, deCODE genetics, and the Intermountain Health. Briefly, we have updated our latest NAFLD GWAS previously published^5^ following similar methods and using the FinnGen data freeze 8. In the UK Biobank analysis, 3363 MASLD cases were added. We added three other cohorts to our meta-analysis from the MASLD GWAS summary statistics from the study from Anstee, et al.^6^ with the EPoS consortium (559 cases and 945 controls), and the study by Sveinbjornsson, et al.^9^: 785 cases from the Icelandic deCODE genetics study, and 2134 cases from the Intermountain dataset (USA). We performed a fixed-effect GWAS meta-analysis of the seven cohorts using METAL^30^ A random-effect analysis was also conducted when variants showed evidence of heterogeneity. A total of 5.8 million of SNPs present in the seven cohorts with a minor allele frequency equal or above 0.01 were investigated in further analyses.

### Mendelian randomization of liver-expressed genes associated and MASLD

We performed two-sample MR analysis between genes located within known MASLD loci as study exposure and MASLD as the study outcome. Each liver-expressed gene with at least one eQTL genome-wide significant was used as exposure (p-value ≤ 5×10^−8^ and genomic window = ± 500 kbp on either side of the lead SNP). Variants in linkage disequilibrium (R2 > 0.1) were removed to retain only independent variants. Summary statistics of the genetic variants selected of each liver-expressed gene in Quebec Obesity Biobank cohort were used to assess their potential causal effect on MASLD by performing Wald ratio or multiple linear regressions (inverse variance weighted) with the mr function from the TwoSampleMR package in R.^31^ The Wald ratio corresponds to the calculation of the effect of a genetic instrument on the outcome (here, MASLD) divided by its effect on the exposure (here, a liver-expressed gene). The IVW-MR method is the analogue of carrying out a meta-analysis of each Wald ratio and was used when a liver-expressed gene had more than three independant genome-wide significant eQTLs. The Benjamini-Hochberg (false discovery rate, FDR) correction was applied to adjust the significance threshold considering the number of comparisons tested (p-value = 1.34×10^−5^ (0.05/3726 genes)).

### Pairwise conditional and colocalization analysis

PWCoCo was used to perform conditional analyses to identify independent MASLD signals at the 8q24.13 genomic region.^32^ Colocalization of each pair of conditionally independent signals for the two traits (eQTLs and MASLD) was performed using summary-level data, which allows for the stringent single-variant assumption to hold for each pair of colocalization analysis. PWCoCo isolated the signal (conditionally independent eQTLs) which colocalize between the two datasets. Briefly, this framework integrates approximate conditional analyses (as implemented in GCTA-COJO^33^), which systematically conditions on each of the association signals within a genomic region and applies pairwise Bayesian colocalization analyses for each pair of independent signals (as implemented in the coloc R package^34^).

### Tissue-specificity of TRIBIAL expression

The tissue-specific gene expression metric (Tau) was obtained for *TRIB1AL*. We used the formula from Yanai et al.^35^ to compare *TRIB1AL* gene expression level across non-sex-specific tissues based on RNA sequencing data from European ancestry donors from Genotype Tissue Expression GTEx (version 10). The RNA-seq data were first log-transformed. After the normalization, a mean value from all samples for each tissue separately was calculated. A Tau specificity score closer to 1 indicates tissue-specificity while a Tau value closer to 0 indicates ubiquitous gene expression.

### Cardiometabolic effects of TRIB1 and TM6SF2 protein-truncating variants

The study design and population of the UK Biobank have previously been reported.^36^ UK Biobank includes more than 500,000 participants between 40 and 69 years old who were recruited from the United Kingdom’s National Health Service (NHS) central registers between 2006 and 2010. Assessment for diseases and serological tests for these participants were done in 22 assessment centers during the same period. Data collection included a self-report questionnaire, physical measures, and blood, urine, and saliva sample collection. Analyses in UK Biobank were conducted under the data application number 25205. PTVs in *TRIB1* and *TM6SF2* (the latter used as a positive control) were extracted from the 389,471 White British participants with available plasma triglyceride levels and whole-exome sequencing data, as previously described.^37^ Of them 34,341 had information on liver fat accumulation measured by magnetic resonance imaging, as previously described.^38^ PTVs were defined as variants causing the loss of an exon or a frameshift in the translation or if they introduced a stop codon, a splice acceptor or a splice donor site.^39^ In order to confirm that the identified PTVs are considered as strong, we used the Ensembl variant effect predictor (VEP) tool^40^ to extract the Combined Annotation Dependent Depletion (CADD) Phred scores for each variant. We also used gnomAD^41^ to extract the predicted loss-of-function (pLoF) value for each PTV. We considered a PTV to be strong if it had either a CADD Phred score above 20 (in the top 1% most deleterious substitutions of the genome) or a high-confidence pLoF value. Noncarriers were defined as having none of the variants identified. This procedure was performed independently for *TRIB1* and *TM6SF2*. All PTV carriers were grouped together and the impact of carrier status on triglyceride levels and liver fat accumulation was determined using two-sided Welch t-tests.

### Cardiometabolic effects of liver TRIBAL expression

Two-sample MR analyses of liver *TRIB1AL*, *PCSK9*, and *ANGPTL3* expression as exposures and multiple cardiometabolic outcomes were performed using the TwoSampleMR R package.^31^ To proxy the liver *TRIB1AL* expression, we selected the rs28601761 variant, prioritized by our PWCoCo analysis, in the summary statistics of *TRIB1AL* eQTL from Quebec Obesity Biobank cohort. Liver *PCSK9*, and *ANGPTL3* eQTLs were used as positive controls and for comparative purpose. To mimic liver-target PCSK9 RNA interference therapies, we selected the variant rs472495 as previously described by Gagnon et al.^42^ For *ANGPTL3*, the strongest cis-acting eQTL in 1 MB upstream and downstream of the *ANGPTL3* gene region (rs1168040) was selected, as previously described.^12^ Summary statistics from GWAS for cardiometabolic outcomes, all publicly available, were selected based on greatest expected statistical power (highest total sample size and cases/controls ratio). Relevant information on GWAS summary statistics used throughout this study is presented in Supplementary Table 4. Glomerular filtration rate, systolic blood pressure, diastolic blood pressure, fasting insulin, fasting glucose, parental lifespan, lipoprotein(a), alanine aminotransferase, and aspartate aminotransferase were reported in log(eGFR), mmHg, mmHg, μU/mL, mmol/L, years, mg/DL, U/L, and U/L, respectively. To allow comparison between measures, we standardised measurements to have standard deviation (SD) of one. We estimated standard deviation using the function sdY.est from the coloc R package. All other continuous variables were rank-based inverse normal transformed prior to GWAS. The causal estimates of these variants mimicking liver-target RNA interference therapies on these cardiometabolic traits were obtained via the Wald ratio MR method.

### Mendelian randomization of liver TRIBAL expression on the liver transcriptome and proteome

The causal effect of liver *TRIB1AL* expression, instrumentalized with genetic variant rs28601761, on 22,719 liver gene expression levels was obtained via a one-sample MR framework and Wald ratio analysis. The FDR correction was applied to adjust the significance threshold considering the number of comparisons tested (p-value = 2.20×10^−6^ (0.05/22,719 genes)). Liver concentrations of 2985 proteins were measured in 460 liver samples by LC-MS/MS-DIA (Supplementary methods). The causal effect of genetically predicted liver *TRIB1AL* expression (proxied by rs28601761) on 2,985 protein expressions in liver was obtained with Wald ratio analysis with a one-sample MR framework.

### Mendelian randomization of liver TRIBAL expression on the blood proteome

The impact of genetically predicted liver *TRIB1AL* expression on blood proteome was tested using similar methods described above for liver transcriptome and proteome but with blood protein quantitative trait loci instead of liver gene/protein expression levels as study outcomes. The two-sample MR analysis included over 1,500 blood protein levels measured by Olink Explore 3072 proximity extension assay in >50,000 UK Biobank participants.^43^ The Wald ratio of the effect of rs28601761, eQTL of liver *TRIB1AL* expression, on each individual blood protein levels was assessed, and Benjamini-Hochberg procedure (FDR) was applied to adjust the significance threshold to correct for multiple testing.

### Institutional review board approval

Patients of the Quebec Obesity Biobank provided informed consent to participate to this institutional biobank. The study was conducted in accordance with the Declaration of Helsinki and approved by the Institutional Review Board (Ethics Committee) of Institut universitaire de cardiologie et de pneumologie de Québec-Université Laval (IUCPQ-UL) (approval number 2021-3656; date of approval: June 17^th^ 2021). UK Biobank received approval from the British National Health Service, North West - Haydock Research Ethics Committee (16/NW/0274). The analysis in the UK Biobank were performed using data application number 25205. All genotype and phenotype data were collected according to an informed consent obtained at the baseline assessment from all participants.

### Data availability

GWAS summary statistics of our previous genome-wide meta-analysis of NAFLD is available on the GWAS catalog under study accession number : <u>GCST90091033</u>.^5^ GWAS summary statistics for NAFLD of the eMERGE network and the EPoS cohort were downloaded from the GWAS Catalog (Study accession: <u>GCST008468</u> and <u>GCST90011885</u>). GWAS summary statistics of NAFLD for FinnGen were obtained from the <u>FinnGen portal</u>. deCODE genetics and Intermountain GWAS summary statistic for NAFLD are available at https://www.decode.com/summarydata. The data used for the tissue-specificity analyses described were obtained from the GTEx Portal on 05/23/2025.

### Code availability

We used publicly available softwares and packages. The code used for this manuscript will be made available on GitHub at time of publication. The GWAS meta-analysis was performed using the <u>METAL</u> v.2011-03-25 software. Statistical analyses were performed in R v.4.1.3 (2022-03-10) using publicly accessible functions from the <u>TwoSampleMR</u> v.0.5.6, the <u>MendelianRandomization</u> v.0.9.0, the <u>ieugwasr</u> v.0.1.5, the <u>tidyverse</u> v.2.0.0 , the <u>data.table</u> v.1.15.2, the <u>coloc</u> v.5.2.3 R packages. Graphical visualisation of the results were made in R v.4.1.3 (2022-03-10) using publicly accessible functions from the <u>ggplot2</u> v.3.5.0, the <u>ggrepel</u> v.0.9.5, the <u>ggprism</u> v.1.0.4, and the <u>locuscomparer</u> v.1.0.0 R packages. The Q-Q plot was generated using the <u>qqman</u> v.0.1.9 R package.

## Acknowledgements

The authors would like to thank all study participants as well as all investigators of the studies that were used throughout the course of this investigation. We acknowledge the invaluable collaboration of the surgery team, bariatric surgeons, and biobank staff and would like to thank all study participants from the Institut universitaire de cardiologie et de pneumologie de Québec (IUCPQ).

