## Supplementary Table 1 for "The long non-coding RNA *TRIB1AL* links metabolic dysfunction-associated steatotic liver disease, cardiometabolic risk and human lifespan"

**Supplementary methods**

**Proteomic analyses**

*Sample Preparation*

Proteins from liver tissue were extracted by 20 cycles of sonication (1s on / 1s off, power level 3) with a Sonic Dismembrator 550 (Fisher Scientific) in 250 µL of lysis buffer containing 50 mM ammonium bicarbonate, 0.5% sodium deoxycholate, 50 mM DTT, 1 µM pepstatin, Complete EDTA free 1X (Roche). Samples were then centrifuged at 10,000 x g for 15 min at 4°C. Proteins from 200 µL of the supernatant were precipitated with 1 mL of cold acetone and incubated overnight at -30°C. The resulting protein pellet was recovered by centrifugation at 10,000 × g for 15 min and resuspended in 50 mM ammonium bicarbonate containing 1% sodium deoxycholate. Protein concentration was determined using the Bradford assay. A volume corresponding to 5 µg of proteins was heated at 95°C for 5 min for protein denaturation. Disulfide bonds were then reduced by addition of DTT for 30 min at 37°C at a final concentration of 0.2 mM, followed by alkylation with iodoacetamide for 30 min at 37°C in the dark at a final concentration of 0.8 mM. Enzymatic digestion was carried out overnight at 37°C using 100 ng of trypsin (Promega). The reaction was stopped by acidification with 5 µL of 50% formic acid, followed by centrifugation at 4000 x g for 15 min to precipitate the sodium deoxycholate. The supernatant was vacuum-dried and resuspended in 0.1% formic acid. 500 ng of the resulting peptides were loaded onto Evotips (Evosep Biosystems) following the manufacturer's protocol.

*LC-MS/MS Analysis - DIA*

Samples were analyzed by LC-MS/MS (Liquid Chromatography – tandem Mass Spectrometry) using an Evosep One LC system (Evosep Biosystems) equipped with an 8 cm length, 100 µm internal diameter, 1.5 µm particle size capillary column (Evosep EV1109) heated at 45°C. A pre-programmed gradient (60 samples per day, 21 min gradient). The column was in line an Orbitrap Exploris 480 mass spectrometer (Thermo Fisher Scientific) operating in Data Independent Acquisition (DIA). Full MS resolution was set to 120,000 with an Automatic Gain Control (AGC) target of 300% and a maximum injection time (IT) of 22 ms, and a total of 35 DIA windows of 15 m/z with an overlap of 0.1 m/z were used to cover a precursor mass range of 350–875 m/z at a resolution of 15,000 with an AGC target of 800% and a maximum IT of 22 ms. Normalized HCD collision energy was set at 30%.

*Data Processing*

Mass spectrometry raw files were analyzed with the DIA-NN 1.8.1 software (<https://github.com/vdemichev/DiaNN>) [1] for peptide and protein identification and quantification using a predicted spectral library generated from the Uniprot *Homo sapiens* reference proteome (Proteome ID UP000005640; 82,439 entries; version 2023.07). Trypsin/P was specified as the enzyme, with a maximum of two missed cleavages. Up to two variable modifications per peptide (methionine oxidation, and N-terminal acetylation) were allowed, while carbamidomethylation of cysteine was set as a fixed modification. Peptides with lengths of 7 to 30 amino acids were considered. Only 2+ to 5+ charge state precursors within a 350–900 m/z range were included. Fragment mass range was set to 200–1800 m/z. The Match Between Runs (MBR) option was enabled to enhance peptide identification across the samples. Protein intensities were normalized using the MaxLFQ algorithm of diann R package (https://rdrr.io/github/vdemichev/diann-rpackage/) [2] and missing values were imputed by a noise value corresponding to first percentile of all protein intensity values within one sample. Only proteins identified with a maximum than 30% of imputated values across the dataset and a minimum of 2 identified peptides were kept for quantification.

**Supplementary Figure 1. Genome-wide association study of metabolic dysfunction associated steatotic liver disease.** Manhattan plot depicting single-nucleotide polymorphisms (SNPs) associated with metabolic dysfunction associated steatotic liver disease (MASLD) in the GWAS meta-analysis.

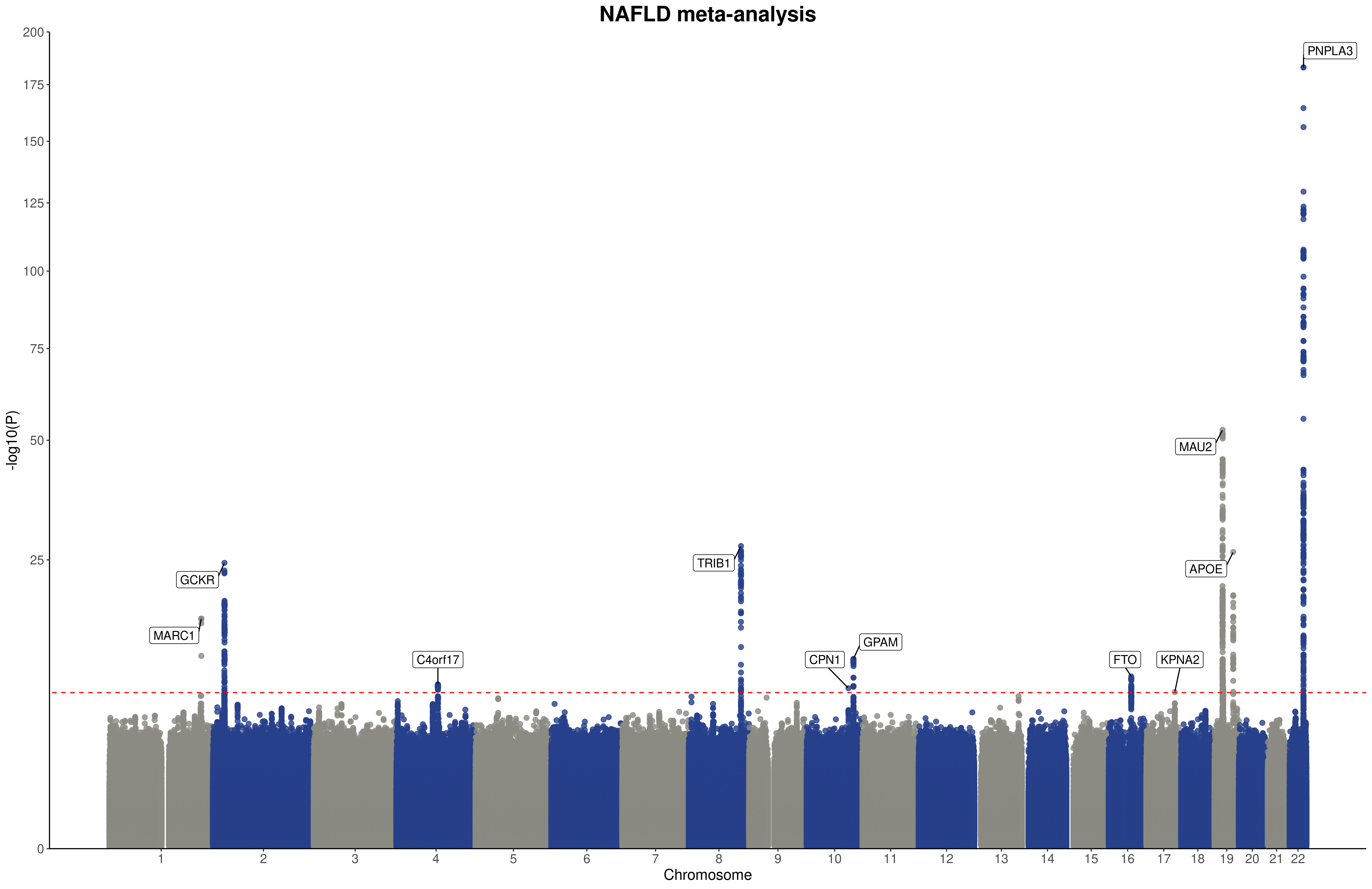

**Supplementary Figure 2. Shared genetic etiology at the *TRIB1AL* locus.** Locuszoom plot depicting colocalization of the top SNP associated with liver *TRIB1AL* expression and metabolic dysfunction associated steatotic liver disease (MASLD). Each dot represents a single-nucleotide polymorphism (SNP) at the *TRIB1AL* locus. These SNPs are plotted to represent their associations with MASLD (top), liver TRIB1 expression (middle), and liver *TRIB1AL* expression (bottom).

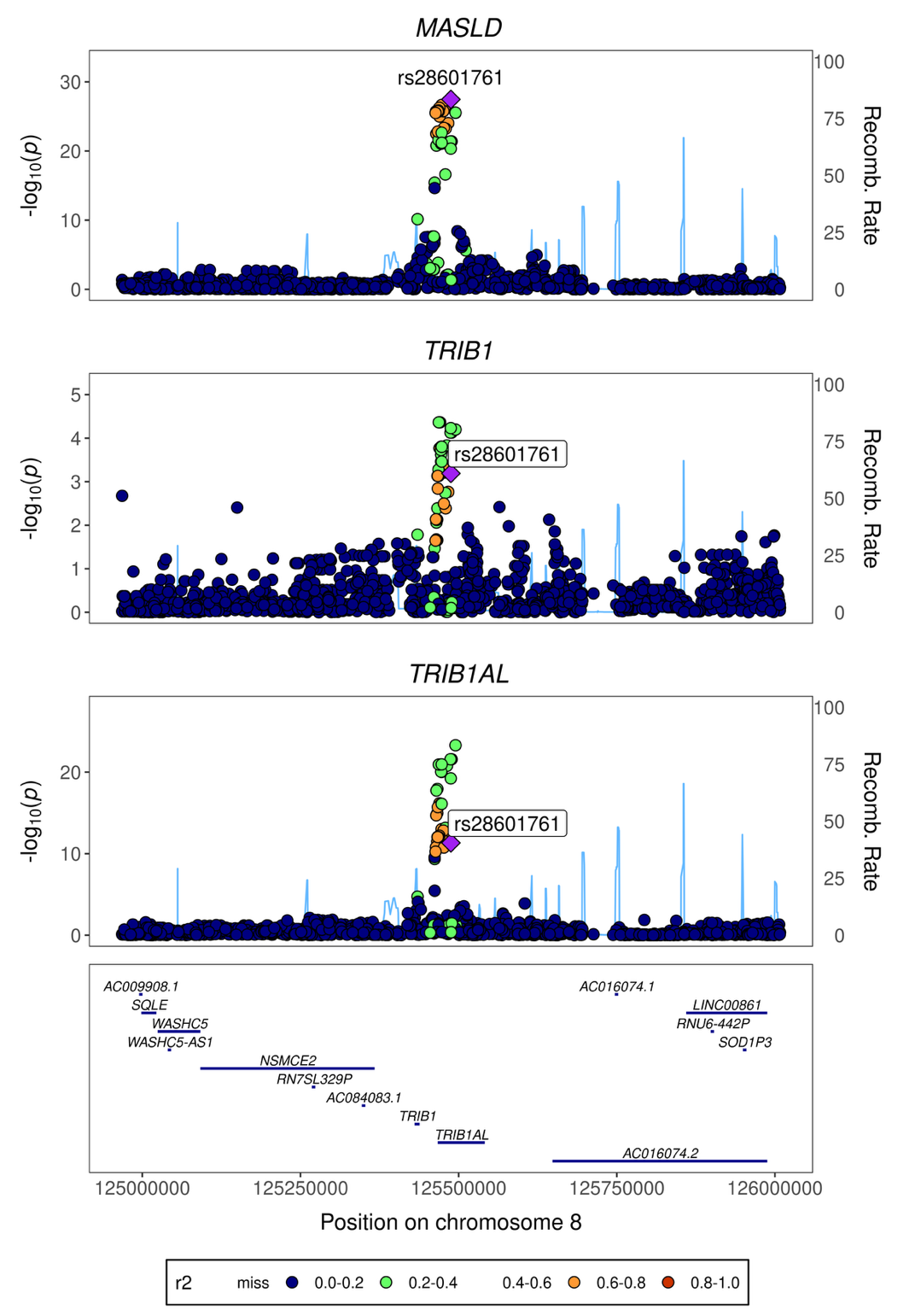

**Supplementary Figure 3. Impact of the causal variant at the *TRIB1* locus on liver *TRIB1AL* expression in participants of the Quebec Obesity Biobank according to stage of liver disease.** rs28601761 was associated with *TRIB1AL* expression levels in A) patients without steatosis, B) in patients with steatosis withous steatohepatitis and C) in patients with steatosis and steatohepatitis. A total of 504 participants of the Quebec Obesity Biobank were separated according to rs28601761 genotype. Gene expression levels of liver *TRIB1AL* were compared using Student t-tests.

A)

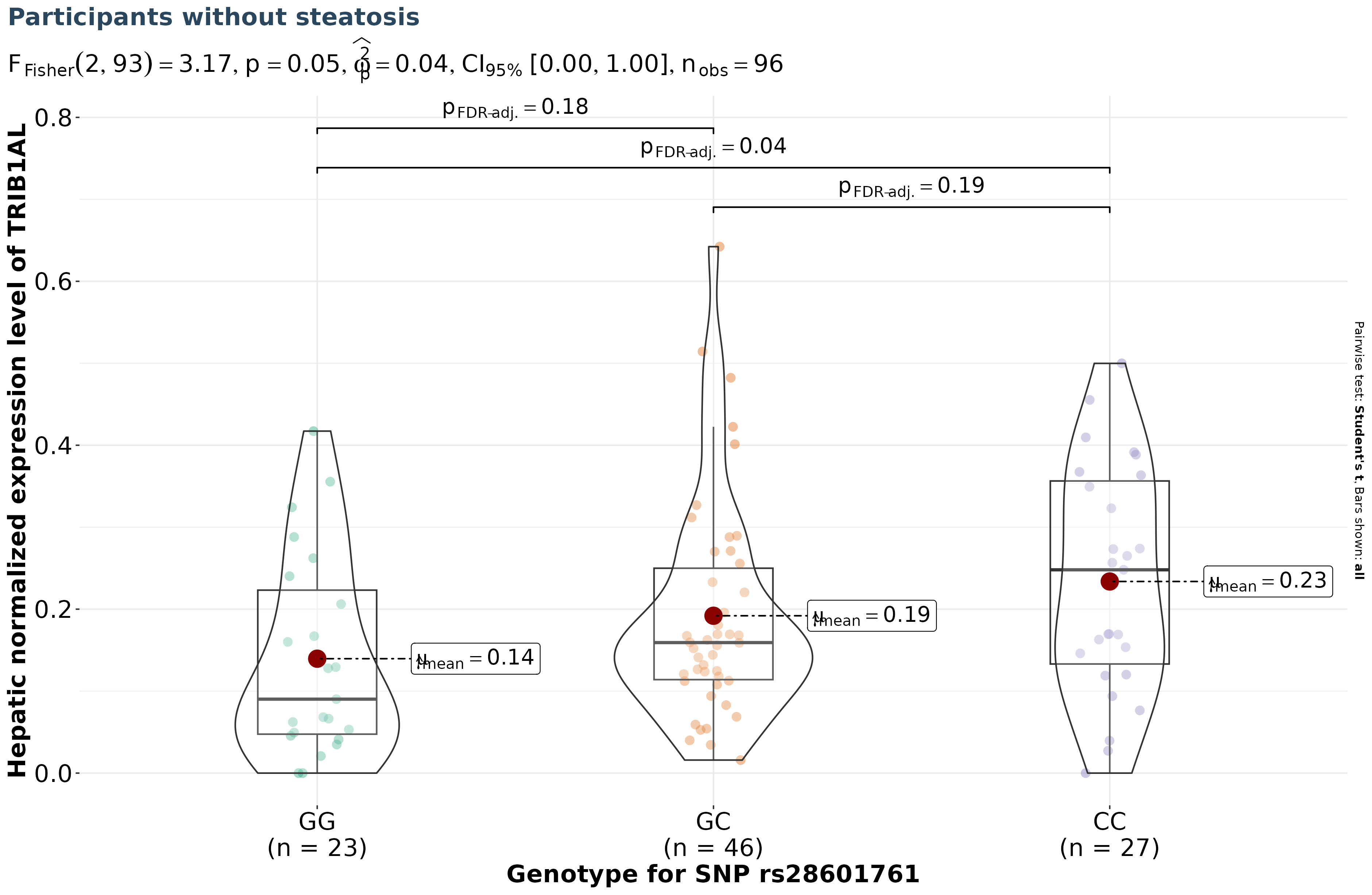

B)

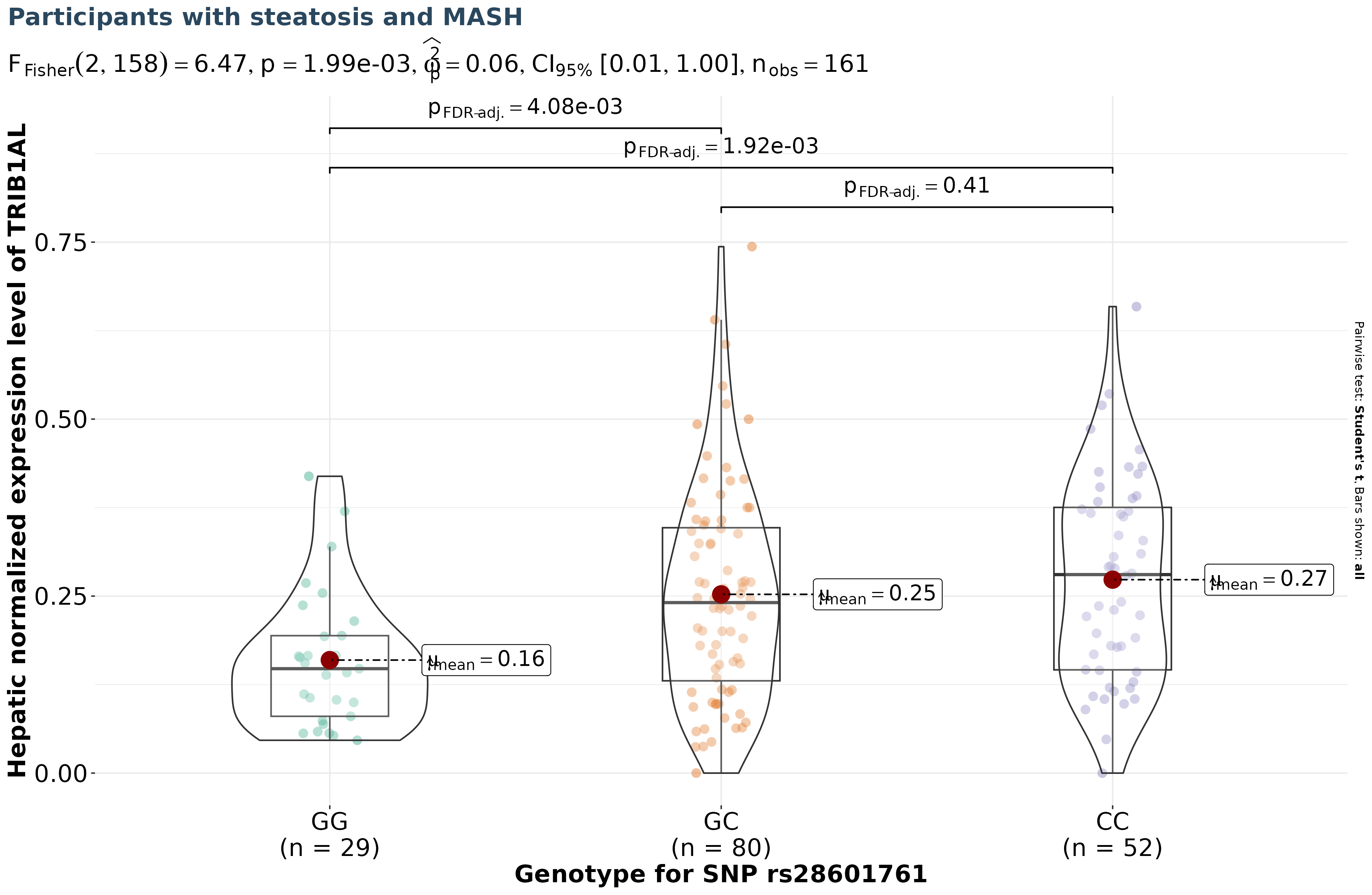

C)

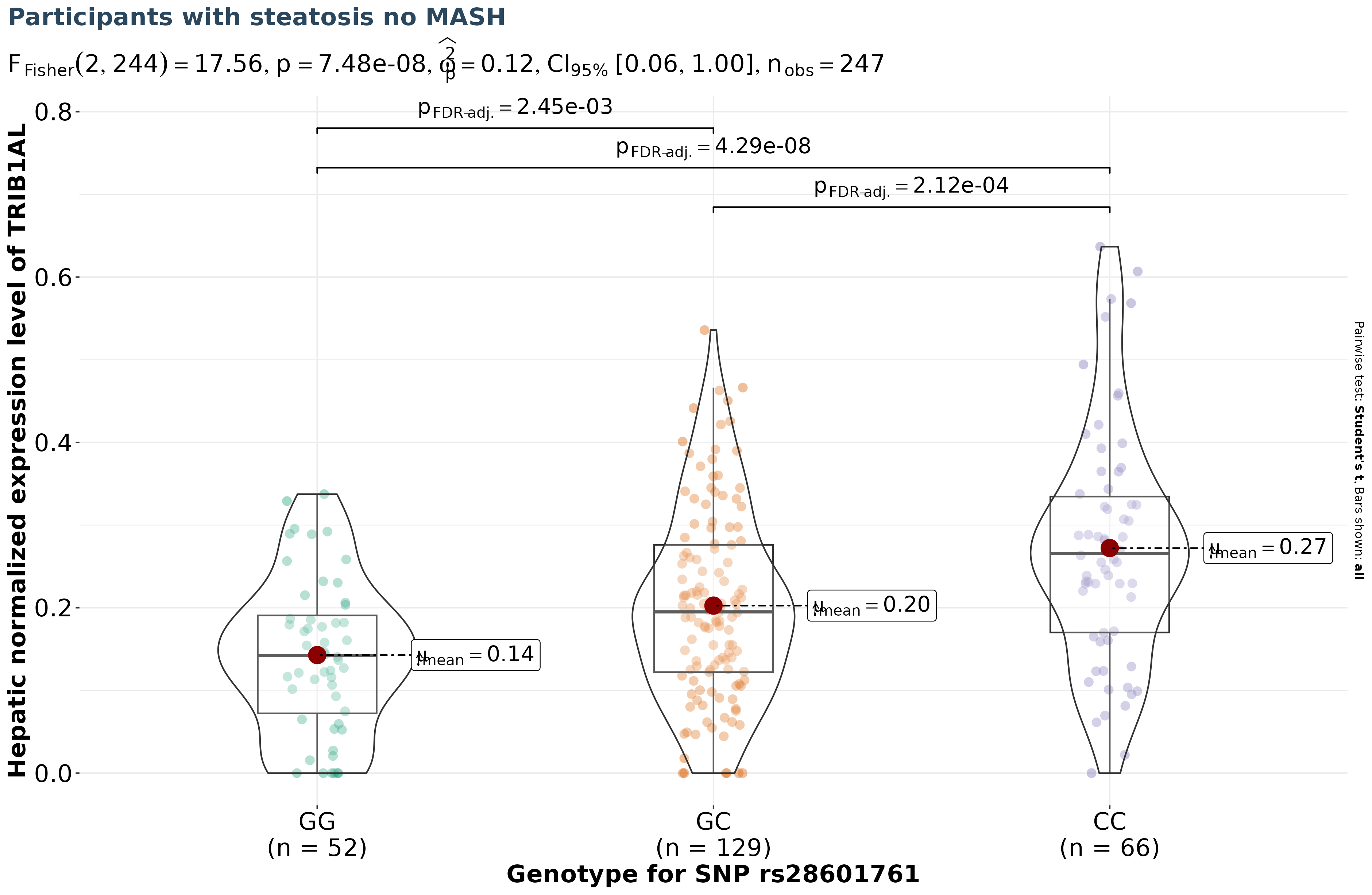

**Supplementary Figure 4. Heat map showing the tissue-specificity of *TRIB1AL*.** Tau value is shown in parentheses after the gene name. Data are presented as averaged log2-transformed transcript by millions (TPM). A Tau specificity score closer to 1 indicates tissue-specificity while a Tau value closer to 0 indicates ubiquitous gene expression.

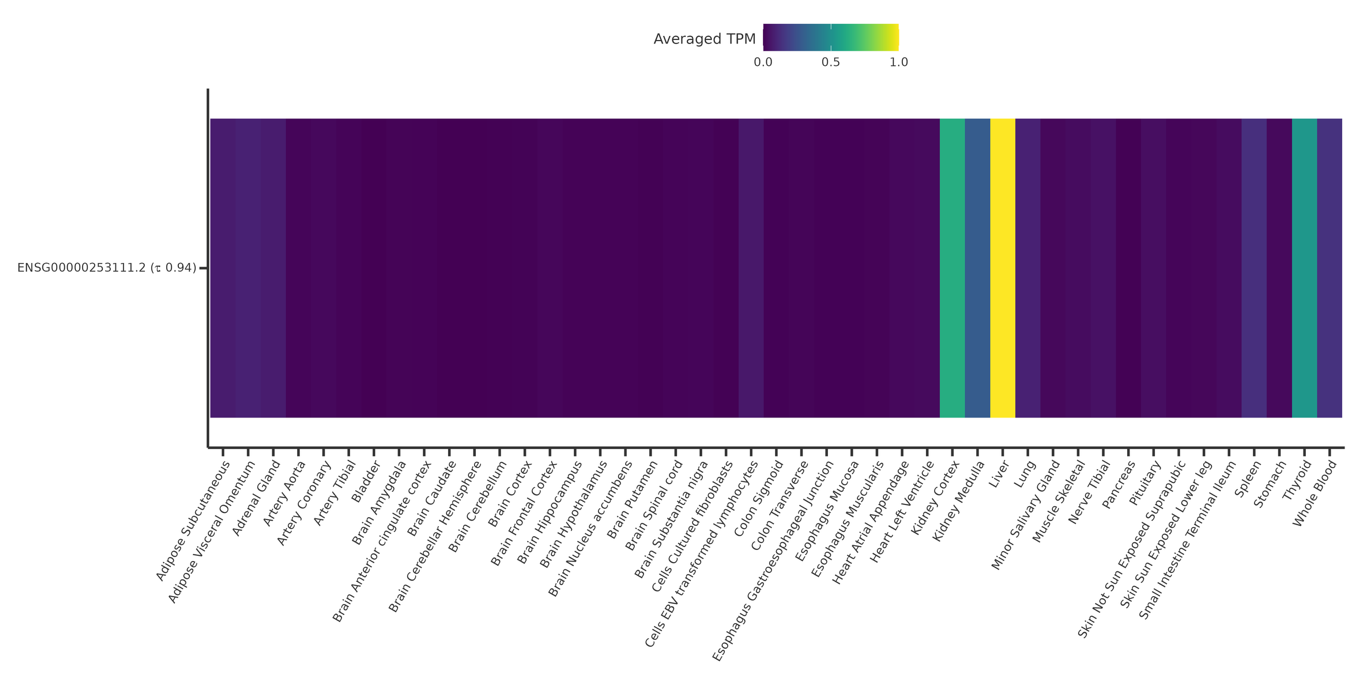

**Supplementary Figure 4. Phenome-wide association study of genetically predicted lower liver TRIB1AL expression.** Genetic association results of variant rs28601761 (8:125487789:C:G) mimicking *TRIB1AL* inhibition on 330 disease endpoints/phecodes from VA Million Veteran Program (3 analyses: n EUR=449,042; n AFR=121,177; n AMR=59,048), FinnGen freeze 12 (n=500,349), and UKBB (pan-UKBB European subset n=420,531). Results were obtained from FinnGen web browser (<https://mvp-ukbb.finngen.fi/variant/8:125487789-C-G>), and represent a multi-way meta-analysis (using standard inverse variance weighting). Triangle pointing downwards indicates that genetic inhibition is associated with lower odds of disease and triangle pointing upwards indicates that genetic inhibition is associated with higher odds of disease,

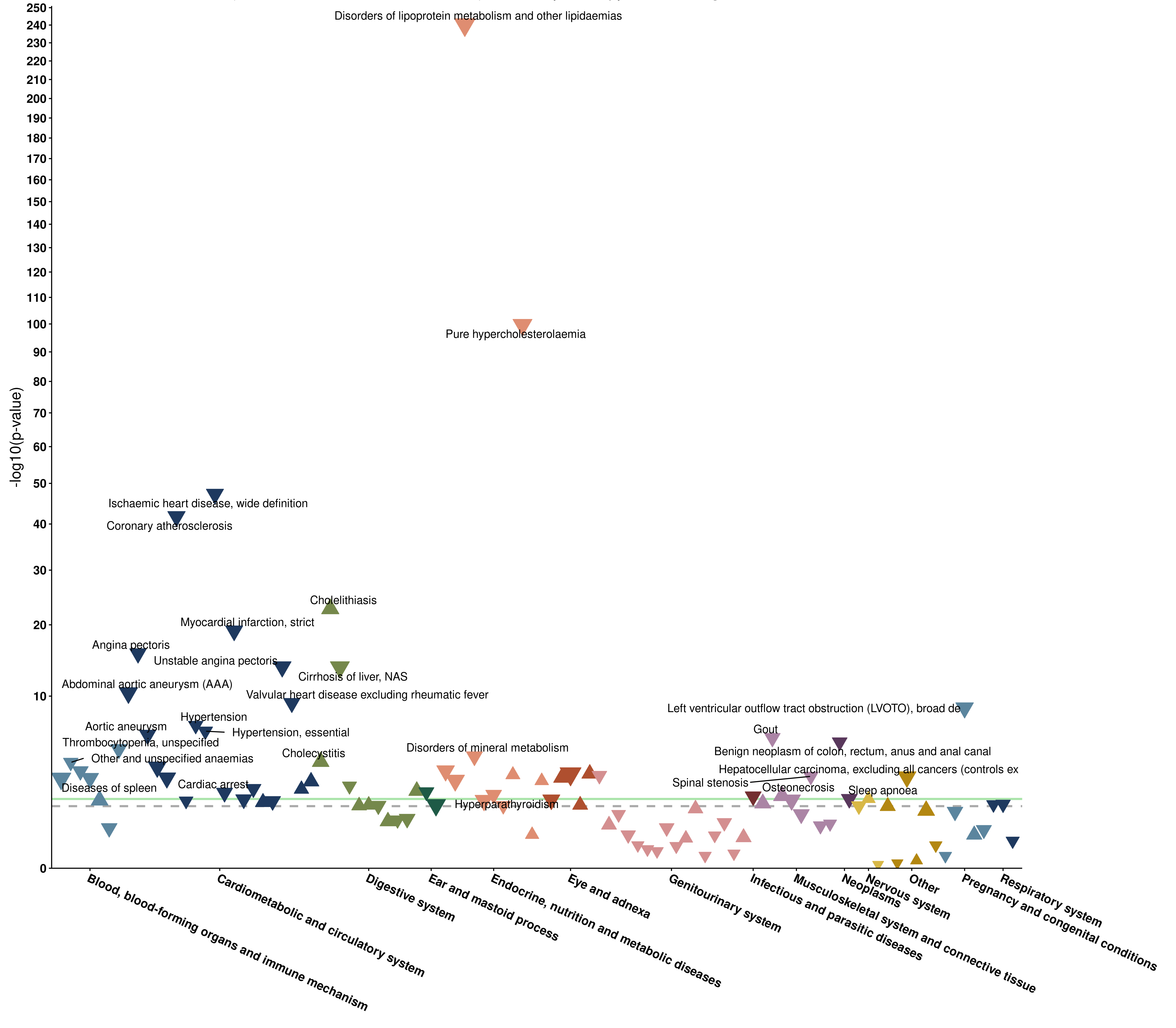

**Supplementary Table 2. Clinical characteristics of the Quebec Obesity Biobank participants.**

| **Characteristic** | **N = 504^1^** |
| --- | --- |
| Female, n (%) | 382 (76%) |
| Age at recruitment | 43 (36, 52) |
| Body mass index, kg/m² | 46 (43, 51) |
| Waist-to-hip ratio | 0.94 (0.88, 1.01) |
| Glucose, mmol/L | 5.80 (5.30, 6.65) |
| Total cholesterol, mmol/L | 4.48 (3.91, 5.11) |
| HDL cholesterol, mmol/L | 1.19 (1.02, 1.40) |
| LDL cholesterol, mmol/L | 2.54 (2.03, 3.13) |
| Triglyceride, mmol/L | 1.44 (1.07, 1.93) |
| Type 2 diabetes, n (%) | 223 (44%) |
| Hypertension, n (%) | 208 (41%) |
| Dyslipidemia, n (%) | 137 (27%) |
| Insulin, pmol/L | 165 (114, 231) |
| Apolipoprotein B, g/L | 0.90 (0.76, 1.07) |
| Hypertension medication, n (%) | 190 (38%) |
| Diabetes medication, n (%) | 93 (18%) |
| Cholesterol medication, n (%) | 119 (24%) |
| Systolic blood pressure, mmHg | 130 (120, 141) |
| Diastolic blood pressure, mmHg | 81 (75, 86) |
| MASLD group, n (%) |  |
| no steatosis | 96 (19%) |
| steatosis no MASH | 247 (49%) |
| steatosis with MASH | 161 (32%) |
| ^1^n (%); Median (Q1, Q3) | |

**Supplementary Table . Cohorts used for the MASLD GWAS and MASLD definitions.**

| **Cohort** | **PMID** | **Accession** | **Population** | **Case** | **Control** | **MASLD definition*** |
| --- | --- | --- | --- | --- | --- | --- |
| deCODE genetics | 36280732 | <https://www.decode.com/summarydata> | Icelandic | 785 | 154465 | ICD-10 code K76.0 (nonalcoholic fatty liver disease) |
| FinnGen study df.8 | <https://finngen.gitbook.io/documentation> | <https://www.finngen.fi/en/access_results> | Finnish | 1908 | 340591 | EHR code K76.0 |
| UK Biobank | 36280732 | <https://www.decode.com/summarydata> | British | 5921 | 402737 | ICD10: K74.0 and K74.2 (hepatic fibrosis), K75.8 (NASH), K76.0 (NAFLD) and ICD10: K76.9 (other specified diseases of the liver) |
| Estonian Biobank | 34841290 | [GCST90091033](https://www.ebi.ac.uk/gwas/studies/GCST90091033) | Estonian | 4119 | 190120 | ICD10: K74.0 and K74.2 (hepatic fibrosis), K75.8 (NASH), K76.0 (NAFLD) and ICD10: K76.9 (other specified diseases of the liver) |
| Intermountain | 36280732 | <https://www.decode.com/summarydata> | American | 2134 | 142760 | ICD-10 code K76.0 (nonalcoholic fatty liver disease) |
| eMERGE Network | 31311600 | GCST008468 | American | 1106 | 8571 | ICD9: 571.5, ICD9: 571.8, ICD9: 571.9, ICD10: K75.81, ICD10: K76.0 and ICD10: K76.9) |
| EPoS Consortium | 32298765 | GCST90011885 | Italian | 559 | 945 | All cases were unrelated patients that had undergone liver biopsy as part of their diagnostic workup for presumed NAFLD. All were originally identified as having abnormal biochemical tests (ALT and/or GGT) and/or an ultrasonographically detected bright liver, associated with features of the metabolic syndrome; or having abnormal biochemical tests (ALT and/or GGT) and macroscopic appearances of a steatotic liver at the time of bariatric surgery. |

*Exclusion criteria included, but were not limited to alcohol dependence, alcoholic liver disease, alpha-1 antitrypsin deficiency, Alagille syndrome, liver transplant, cystic fibrosis, hepatitis, abetalipoproteinemia, LCAT deficiency, lipodystrophy, disorders of copper metabolism Reye’s syndrome, inborn errors of metabolism, HELLP syndrome, starvation and acute fatty liver (as suggested by the American Association for the Study of Liver Disease [AASLD]).

**Supplementary Table 3. Effect of genetically predicted liver expression of genes and known MASLD loci on MASLD.** Study exposures were derived from RNA sequencing of 504 liver samples of the Quebec Obesity Biobank and the study outcome was derived from a new genome-wide meta-analysis of MASLD based on electronic health records. Genetic loci without eQTL available are listed at the end of the table.

| **Exposure** | **Outcome** | **MR method** | **Nsnp** | **Effect size** | **Standard error** | **Pval** |
| --- | --- | --- | --- | --- | --- | --- |
| **TRIB1AL** | MASLD | Wald ratio | 1 | 0.213 | 0.020 | 2.85E-26 |
| **PNPLA3** | MASLD | Wald ratio | 1 | -0.622 | 0.096 | 9.13E-11 |
| **TM6SF2** | MASLD | Wald ratio | 1 | -0.357 | 0.056 | 2.11E-10 |
| **MTTP** | MASLD | Wald ratio | 1 | -0.501 | 0.087 | 1.03E-08 |
| **AKNA** | MASLD | Wald ratio | 1 | 0.146 | 0.030 | 1.03E-06 |
| **EPHA2** | MASLD | Wald ratio | 1 | -0.099 | 0.023 | 1.36E-05 |
| **GPAM** | MASLD | Wald ratio | 1 | 0.253 | 0.063 | 5.32E-05 |
| **CHEK2** | MASLD | Wald ratio | 1 | -0.031 | 0.011 | 3.14E-03 |
| **PPP1R3B** | MASLD | Wald ratio | 1 | 0.094 | 0.033 | 4.34E-03 |
| **PCCB** | MASLD | Wald ratio | 1 | -0.121 | 0.044 | 5.51E-03 |
| **APOH** | MASLD | Wald ratio | 1 | 0.214 | 0.078 | 5.86E-03 |
| **COBLL1** | MASLD | Wald ratio | 1 | -0.283 | 0.104 | 6.80E-03 |
| **BCL7B** | MASLD | Wald ratio | 1 | 0.108 | 0.047 | 0.021 |
| **APOE** | MASLD | Inverse variance weighted | 2 | 0.171 | 0.076 | 0.025 |
| **OSGIN1** | MASLD | Wald ratio | 1 | 0.072 | 0.033 | 0.029 |
| **DHODH** | MASLD | Wald ratio | 1 | 0.127 | 0.058 | 0.030 |
| **HLA-DQA2** | MASLD | Wald ratio | 1 | 0.023 | 0.011 | 0.032 |
| **CD276** | MASLD | Wald ratio | 1 | 0.026 | 0.014 | 0.057 |
| **EFHD1** | MASLD | Wald ratio | 1 | 0.023 | 0.012 | 0.058 |
| **FADS2** | MASLD | Wald ratio | 1 | -0.068 | 0.038 | 0.069 |
| **IL1R2** | MASLD | Wald ratio | 1 | -0.040 | 0.022 | 0.073 |
| **HKDC1** | MASLD | Wald ratio | 1 | 0.023 | 0.014 | 0.090 |
| **ABO** | MASLD | Wald ratio | 1 | 0.025 | 0.015 | 0.102 |
| **TMEM147** | MASLD | Wald ratio | 1 | -0.018 | 0.011 | 0.106 |
| **MLXIP** | MASLD | Wald ratio | 1 | -0.066 | 0.041 | 0.107 |
| **NYNRIN** | MASLD | Wald ratio | 1 | 0.019 | 0.012 | 0.123 |
| **SLC2A2** | MASLD | Inverse variance weighted | 2 | -0.162 | 0.108 | 0.133 |
| **P2RX7** | MASLD | Wald ratio | 1 | -0.035 | 0.028 | 0.211 |
| **ERLIN1** | MASLD | Wald ratio | 1 | -0.080 | 0.066 | 0.222 |
| **APOL3** | MASLD | Wald ratio | 1 | -0.056 | 0.048 | 0.241 |
| **PIGV** | MASLD | Wald ratio | 1 | -0.082 | 0.072 | 0.253 |
| **STAP2** | MASLD | Wald ratio | 1 | 0.027 | 0.025 | 0.286 |
| **MLYCD** | MASLD | Wald ratio | 1 | 0.058 | 0.057 | 0.311 |
| **HP** | MASLD | Wald ratio | 1 | 0.035 | 0.037 | 0.333 |
| **OGFRL1** | MASLD | Wald ratio | 1 | -0.059 | 0.064 | 0.357 |
| **DNAJC22** | MASLD | Wald ratio | 1 | 0.026 | 0.029 | 0.368 |
| **FCGR2B** | MASLD | Wald ratio | 1 | 0.023 | 0.029 | 0.427 |
| **CPS1** | MASLD | Wald ratio | 1 | 0.057 | 0.073 | 0.432 |
| **NRXN2** | MASLD | Wald ratio | 1 | 0.010 | 0.013 | 0.435 |
| **FTO** | MASLD | Wald ratio | 1 | -0.047 | 0.068 | 0.487 |
| **PANX1** | MASLD | Wald ratio | 1 | 0.034 | 0.051 | 0.505 |
| **MPND** | MASLD | Wald ratio | 1 | 0.032 | 0.049 | 0.518 |
| **TNFSF10** | MASLD | Wald ratio | 1 | -0.028 | 0.046 | 0.545 |
| **TRPS1** | MASLD | Wald ratio | 1 | 0.014 | 0.028 | 0.611 |
| **PRAG1** | MASLD | Wald ratio | 1 | -0.021 | 0.047 | 0.660 |
| **S1PR2** | MASLD | Wald ratio | 1 | 0.008 | 0.020 | 0.694 |
| **SHROOM3** | MASLD | Wald ratio | 1 | 0.010 | 0.026 | 0.707 |
| **TRIM5** | MASLD | Inverse variance weighted | 2 | 0.012 | 0.037 | 0.754 |
| **PAQR9** | MASLD | Wald ratio | 1 | 0.006 | 0.021 | 0.775 |
| **IL1RN** | MASLD | Wald ratio | 1 | -0.015 | 0.054 | 0.778 |
| **ITCH** | MASLD | Wald ratio | 1 | -0.019 | 0.067 | 0.780 |
| **SOX7** | MASLD | Inverse variance weighted | 2 | -0.005 | 0.022 | 0.811 |
| **RORA** | MASLD | Wald ratio | 1 | 0.024 | 0.116 | 0.838 |
| **MERTK** | MASLD | Inverse variance weighted | 2 | 0.005 | 0.027 | 0.843 |
| **SMARCD2** | MASLD | Wald ratio | 1 | 0.007 | 0.043 | 0.879 |
| **DDX42** | MASLD | Wald ratio | 1 | -0.004 | 0.042 | 0.921 |
| **SCN2A** | MASLD | Wald ratio | 1 | 0.003 | 0.037 | 0.940 |
| **ANPEP** | MASLD | Wald ratio | 1 | 0.001 | 0.021 | 0.969 |
| **CASP8** | MASLD | Wald ratio | 1 | 0.000 | 0.042 | 0.992 |
| **GPN2** |  |  |  |  |  |  |
| **KDF1** |  |  |  |  |  |  |
| **SYTL1** |  |  |  |  |  |  |
| **CCDC18** |  |  |  |  |  |  |
| **FCGR2A** |  |  |  |  |  |  |
| **SLC30A10** |  |  |  |  |  |  |
| **MTARC1** |  |  |  |  |  |  |
| **CRIM1** |  |  |  |  |  |  |
| **TBC1D8** |  |  |  |  |  |  |
| **ABCB11** |  |  |  |  |  |  |
| **lnc-RHBDD1** |  |  |  |  |  |  |
| **PPARG** |  |  |  |  |  |  |
| **U2SURP** |  |  |  |  |  |  |
| **TM4SF1** |  |  |  |  |  |  |
| **HSD17B13** |  |  |  |  |  |  |
| **MAP3K1** |  |  |  |  |  |  |
| **MLXIPL** |  |  |  |  |  |  |
| **ABCB4** |  |  |  |  |  |  |
| **KLF10** |  |  |  |  |  |  |
| **WASHC5** |  |  |  |  |  |  |
| **GPT** |  |  |  |  |  |  |
| **UHRF2** |  |  |  |  |  |  |
| **TJP2** |  |  |  |  |  |  |
| **PSD** |  |  |  |  |  |  |
| **FBXL15** |  |  |  |  |  |  |
| **CUX2** |  |  |  |  |  |  |
| **SH2B3** |  |  |  |  |  |  |
| **ATXN2** |  |  |  |  |  |  |
| **HNF1A** |  |  |  |  |  |  |
| **FLT1** |  |  |  |  |  |  |
| **SERPINA1** |  |  |  |  |  |  |
| **KAT7** |  |  |  |  |  |  |
| **NEDD4L** |  |  |  |  |  |  |
| **PIK3R2** |  |  |  |  |  |  |
| **IFI30** |  |  |  |  |  |  |
| **MPV17L2** |  |  |  |  |  |  |
| **CEBPA** |  |  |  |  |  |  |
| **CEBPG** |  |  |  |  |  |  |
| **MAFB** |  |  |  |  |  |  |

**Supplementary Table 4. Characteristics of the genome-wide association studies used for Mendelian randomization analyses.**

| **Phenotype** | **Year** | **PMID** | **Consortium** | **Population** | **Sample size** | **ncase** | **ncontrol** |
| --- | --- | --- | --- | --- | --- | --- | --- |
| Abdominal subcutaneous adipose tissue (ASAT) | 2022 | 35773277 | UK Biobank (UKB) | Mixed | 39076 | - | - |
| Visceral adipose tissue (VAT) | 2022 | 35773277 | UKB | Mixed | 39076 | - | - |
| Diastolic blood pressure | 2018 | 30224653 | International Consortium of Blood Pressure | European | 757601 | - | - |
| Systolic blood pressure | 2018 | 30224653 | International Consortium of Blood Pressure | European | 757601 | - | - |
| Fasting glucose | 2021 | 33402679 | MAGIC | European | 151188 | - | - |
| Fasting insulin | 2021 | 33402679 | MAGIC | European | 105056 | - | - |
| Glomerular filtration rate | 2021 | 34272381 | CKDGEN, UKB | European | 1004040 | - | - |
| Healthspan | 2019 | 30729179 | UKB | European | 300447 | - | - |
| Parental lifespan | 2019 | 30642433 | UKB, LifeGen | European | 253060 | - | - |
| Apolipoprotein B | 2020 | 32203549 | UKB | European | 439214 | - | - |
| HDL cholesterol | 2021 | 34887591 | GLGC | Mixed | 1547630 | - | - |
| LDL cholesterol | 2021 | 34887591 | GLGC | Mixed | 1494170 | - | - |
| Lipoprotein (a) | 2021 | 33462484 | UKB | European | 363228 | - | - |
| Triglyceride | 2021 | 34887591 | GLGC | Mixed | 1521780 | - | - |
| Alanine aminotransferase | 2021 | 34315874 | UKB | European | 387859 | - | - |
| Aspartate aminotransferase | 2021 | 34315874 | UKB | European | 386570 | - | - |
| Liver Fat | 2021 | 34128465 | UKB | European | 38000 | - | - |
| Calcific aortic valve stenosis | 2024 | 38494474 | Six cohorts | European | 941863 | 14819 | 927044 |
| Coronary artery disease | 2022 | 36474045 | UKB+CARDIoGRAMplusC4D+nine studies | European | 1347212 | 181522 | 1165690 |
| Heart failure | 2020 | 31919418 | HERMES | European | 977323 | 47309 | 930014 |
| Ischemic stroke | 2022 | 36180795 | UKB, ISGC | European | 1614080 | 62100 | 1234808 |
| Type 2 diabetes | 2022 | 35551307 | UKB, DIAMANTE | European | 933970 | 80154 | 853816 |
| Chronic Kidney disease | 2019 | 31152163 | CKDGen | European | 480698 | 41395 | 439303 |
| Longevity | 2019 | 31413261 | ±20 cohorts | European | 36745 | 11262 | 25483 |
| MASLD | 2024 | 34841290  36280732 | UKB, FinnGen study df.8, Estonian Biobank, deCODE genetics, Intermountain, eMERGE Network, EPoS Consortium | European | 1256720 | 16532 | 1240188 |

**Supplementary Table 5. PTV at *TRIB1* with high impact or missense variants and CADD score ≥ 20.**

| **Variant ID** | **RSID** | **Consequence (Most severe by gene)** | **Population AF** |
| --- | --- | --- | --- |
| **8_125430907_G_A** | rs988507358 | [""""missense_variant:TRIB1""""] | 3.19E-06 |
| **8_125430916_C_T** |  | [""""missense_variant:TRIB1""""] | 2.13E-06 |
| **8_125430922_G_A** | rs1266069049 | [""""missense_variant:TRIB1""""] | 1.06E-06 |
| **8_125430925_C_G** |  | [""""missense_variant:TRIB1""""] | 3.19E-06 |
| **8_125430928_C_T** | rs1176986805 | [""""missense_variant:TRIB1""""] | 5.32E-06 |
| **8_125430931_T_C** |  | [""""missense_variant:TRIB1""""] | 4.26E-06 |
| **8_125430932_G_A** |  | [""""missense_variant:TRIB1""""] | 3.19E-06 |
| **8_125430936_G_A** | rs1440081687 | [""""missense_variant:TRIB1""""] | 9.58E-06 |
| **8_125430936_G_T** |  | [""""missense_variant:TRIB1""""] | 1.06E-06 |
| **8_125430943_C_T** |  | [""""missense_variant:TRIB1""""] | 2.13E-06 |
| **8_125430955_G_A** |  | [""""missense_variant:TRIB1""""] | 1.06E-06 |
| **8_125430955_G_T** | rs747946279 | [""""missense_variant:TRIB1""""] | 5.76E-04 |
| **8_125430957_C_T** | rs1390586417 | [""""missense_variant:TRIB1""""] | 3.19E-06 |
| **8_125430967_T_A** |  | [""""missense_variant:TRIB1""""] | 4.26E-06 |
| **8_125430973_C_T** |  | [""""missense_variant:TRIB1""""] | 3.19E-05 |
| **8_125430982_G_A** | rs1039513360 | [""""missense_variant:TRIB1""""] | 2.13E-06 |
| **8_125430999_C_T** |  | [""""missense_variant:TRIB1""""] | 5.32E-06 |
| **8_125431014_G_A** |  | [""""missense_variant:TRIB1""""] | 1.06E-06 |
| **8_125431032_G_C** |  | [""""missense_variant:TRIB1""""] | 8.51E-06 |
| **8_125431033_C_A** | rs1235489441 | [""""missense_variant:TRIB1""""] | 3.19E-06 |
| **8_125431035_G_A** |  | [""""missense_variant:TRIB1""""] | 2.13E-06 |
| **8_125431038_A_C** | rs1208735184 | [""""missense_variant:TRIB1""""] | 4.26E-06 |
| **8_125431044_C_T** | rs1563823407 | [""""missense_variant:TRIB1""""] | 5.32E-06 |
| **8_125431047_C_T** | rs1415420971 | [""""missense_variant:TRIB1""""] | 1.06E-06 |
| **8_125431051_T_A** |  | [""""missense_variant:TRIB1""""] | 2.13E-06 |
| **8_125431057_A_G** |  | [""""missense_variant:TRIB1""""] | 9.58E-06 |
| **8_125431058_G_C** |  | [""""missense_variant:TRIB1""""] | 1.06E-06 |
| **8_125431071_C_T** |  | [""""missense_variant:TRIB1""""] | 1.06E-06 |
| **8_125431090_C_A** | rs1448026346 | [""""missense_variant:TRIB1""""] | 2.13E-06 |
| **8_125431090_C_T** | rs1448026346 | [""""missense_variant:TRIB1""""] | 1.70E-05 |
| **8_125431092_G_C** | rs1402829800 | [""""missense_variant:TRIB1""""] | 1.06E-06 |
| **8_125431108_C_T** | rs1001989436 | [""""missense_variant:TRIB1""""] | 2.02E-05 |
| **8_125431113_C_A** |  | [""""missense_variant:TRIB1""""] | 1.06E-06 |
| **8_125431113_C_T** | rs1472661390 | [""""missense_variant:TRIB1""""] | 1.06E-06 |
| **8_125431149_G_A** | rs1399514169 | [""""missense_variant:TRIB1""""] | 1.06E-06 |
| **8_125431153_G_T** | rs765931219 | [""""missense_variant:TRIB1""""] | 1.58E-04 |
| **8_125431162_G_A** | rs895790586 | [""""missense_variant:TRIB1""""] | 2.66E-05 |
| **8_125431168_G_A** |  | [""""missense_variant:TRIB1""""] | 1.06E-06 |
| **8_125431171_G_T** |  | [""""missense_variant:TRIB1""""] | 5.32E-06 |
| **8_125431176_G_T** | rs766476837 | [""""missense_variant:TRIB1""""] | 6.39E-06 |
| **8_125431179_G_C** |  | [""""missense_variant:TRIB1""""] | 7.45E-06 |
| **8_125431186_T_C** |  | [""""missense_variant:TRIB1""""] | 1.06E-06 |
| **8_125431201_C_T** | rs751760974 | [""""missense_variant:TRIB1""""] | 9.58E-06 |
| **8_125431215_G_C** |  | [""""missense_variant:TRIB1""""] | 1.06E-06 |
| **8_125431216_T_C** |  | [""""missense_variant:TRIB1""""] | 5.32E-06 |
| **8_125431235_C_G** |  | [""""missense_variant:TRIB1""""] | 1.28E-05 |
| **8_125431240_C_A** |  | [""""missense_variant:TRIB1""""] | 3.19E-06 |
| **8_125431240_C_T** |  | [""""missense_variant:TRIB1""""] | 1.06E-06 |
| **8_125431254_C_A** | rs1026856398 | [""""missense_variant:TRIB1""""] | 1.28E-05 |
| **8_125431255_G_A** |  | [""""missense_variant:TRIB1""""] | 2.13E-06 |
| **8_125431255_G_T** |  | [""""missense_variant:TRIB1""""] | 1.06E-06 |
| **8_125433335_T_C** | rs201548983 | [""""missense_variant:TRIB1""""] | 1.38E-05 |
| **8_125433357_A_G** | rs750474120 | [""""missense_variant:TRIB1""""] | 2.13E-05 |
| **8_125433372_C_T** | rs1457004284 | [""""missense_variant:TRIB1""""] | 3.19E-06 |
| **8_125433381_A_C** |  | [""""missense_variant:TRIB1""""] | 3.19E-06 |
| **8_125433382_C_A** | rs749750278 | [""""missense_variant:TRIB1""""] | 1.81E-05 |
| **8_125433383_A_G** | rs1239143332 | [""""missense_variant:TRIB1""""] | 1.06E-06 |
| **8_125433384_T_C** | rs1440108532 | [""""missense_variant:TRIB1""""] | 5.32E-06 |
| **8_125433398_G_C** |  | [""""missense_variant:TRIB1""""] | 8.51E-06 |
| **8_125433402_T_A** | rs200241998 | [""""missense_variant:TRIB1""""] | 1.17E-05 |
| **8_125433411_G_A** |  | [""""missense_variant:TRIB1""""] | 1.06E-06 |
| **8_125433422_G_A** |  | [""""missense_variant:TRIB1""""] | 3.19E-06 |
| **8_125433426_A_G** | rs779298762 | [""""missense_variant:TRIB1""""] | 3.19E-06 |
| **8_125433429_T_A** |  | [""""missense_variant:TRIB1""""] | 4.26E-06 |
| **8_125433431_T_G** |  | [""""missense_variant:TRIB1""""] | 1.06E-06 |
| **8_125433434_T_C** |  | [""""missense_variant:TRIB1""""] | 2.13E-06 |
| **8_125433438_A_G** |  | [""""missense_variant:TRIB1""""] | 2.13E-06 |
| **8_125433446_T_A** |  | [""""missense_variant:TRIB1""""] | 1.06E-06 |
| **8_125433456_T_C** | rs751349356 | [""""start_lost:TRIB1""""] | 5.32E-06 |
| **8_125433471_G_A** | rs1199031481 | [""""missense_variant:TRIB1""""] | 4.26E-06 |
| **8_125433475_C_G** | rs56285697 | [""""missense_variant:TRIB1""""] | 1.06E-05 |
| **8_125433476_C_T** | rs758099027 | [""""missense_variant:TRIB1""""] | 5.11E-05 |
| **8_125433486_T_C** | rs1477651120 | [""""missense_variant:TRIB1""""] | 1.17E-05 |
| **8_125433488_C_T** | rs779326105 | [""""missense_variant:TRIB1""""] | 6.39E-06 |
| **8_125433489_G_A** | rs748516687 | [""""missense_variant:TRIB1""""] | 4.79E-05 |
| **8_125433500_G_A** |  | [""""missense_variant:TRIB1""""] | 3.19E-06 |
| **8_125433503_G_A** | rs778292179 | [""""missense_variant:TRIB1""""] | 4.26E-06 |
| **8_125433506_C_T** | rs371769788 | [""""missense_variant:TRIB1""""] | 1.01E-04 |
| **8_125433507_G_A** | rs150043688 | [""""missense_variant:TRIB1""""] | 4.26E-06 |
| **8_125433515_A_G** | rs754492794 | [""""missense_variant:TRIB1""""] | 1.06E-06 |
| **8_125433530_G_A** | rs149162682 | [""""missense_variant:TRIB1""""] | 7.45E-06 |
| **8_125433533_G_A** | rs140863521 | [""""missense_variant:TRIB1""""] | 9.58E-06 |
| **8_125433537_C_T** |  | [""""missense_variant:TRIB1""""] | 2.13E-06 |
| **8_125433539_C_G** |  | [""""missense_variant:TRIB1""""] | 1.06E-06 |
| **8_125433547_C_G** |  | [""""missense_variant:TRIB1""""] | 1.06E-06 |
| **8_125433548_C_T** |  | [""""stop_gained:TRIB1""""] | 1.06E-06 |
| **8_125433560_G_A** | rs199695896 | [""""missense_variant:TRIB1""""] | 1.09E-04 |
| **8_125433563_C_G** |  | [""""missense_variant:TRIB1""""] | 2.13E-06 |
| **8_125433569_G_A** | rs1197954090 | [""""missense_variant:TRIB1""""] | 3.19E-06 |
| **8_125433589_C_G** |  | [""""missense_variant:TRIB1""""] | 1.06E-06 |
| **8_125433590_G_A** | rs1273942367 | [""""missense_variant:TRIB1""""] | 2.13E-06 |
| **8_125433597_C_G** |  | [""""missense_variant:TRIB1""""] | 4.26E-06 |
| **8_125433602_G_C** |  | [""""missense_variant:TRIB1""""] | 1.06E-06 |
| **8_125433604_G_T** |  | [""""missense_variant:TRIB1""""] | 1.06E-06 |
| **8_125436007_A_C** |  | [""""missense_variant:TRIB1""""] | 1.06E-06 |
| **8_125436023_A_G** | rs747683346 | [""""missense_variant:TRIB1""""] | 2.13E-06 |
| **8_125436046_A_G** |  | [""""start_lost:TRIB1""""] | 2.13E-06 |
| **8_125436053_G_A** | rs147892039 | [""""missense_variant:TRIB1""""] | 5.32E-06 |
| **8_125436053_G_C** | rs147892039 | [""""missense_variant:TRIB1""""] | 1.06E-06 |
| **8_125436056_A_T** |  | [""""missense_variant:TRIB1""""] | 1.06E-06 |
| **8_125436058_G_A** |  | [""""missense_variant:TRIB1""""] | 2.13E-06 |
| **8_125436059_A_G** |  | [""""missense_variant:TRIB1""""] | 1.06E-06 |
| **8_125436064_G_A** |  | [""""missense_variant:TRIB1""""] | 1.06E-06 |
| **8_125436070_T_C** |  | [""""missense_variant:TRIB1""""] | 4.26E-06 |
| **8_125436071_C_T** |  | [""""missense_variant:TRIB1""""] | 2.13E-06 |
| **8_125436073_G_C** |  | [""""missense_variant:TRIB1""""] | 1.06E-06 |
| **8_125436092_C_T** |  | [""""missense_variant:TRIB1""""] | 1.06E-06 |
| **8_125436097_G_A** | rs1257256547 | [""""missense_variant:TRIB1""""] | 2.13E-06 |
| **8_125436101_G_A** | rs1230576914 | [""""missense_variant:TRIB1""""] | 4.26E-06 |
| **8_125436110_T_C** |  | [""""missense_variant:TRIB1""""] | 1.06E-06 |
| **8_125436112_C_G** |  | [""""missense_variant:TRIB1""""] | 6.39E-06 |
| **8_125436125_G_C** |  | [""""missense_variant:TRIB1""""] | 1.06E-06 |
| **8_125436130_T_C** |  | [""""missense_variant:TRIB1""""] | 2.02E-05 |
| **8_125436142_G_A** |  | [""""missense_variant:TRIB1""""] | 1.06E-06 |
| **8_125436146_C_T** | rs1258979472 | [""""missense_variant:TRIB1""""] | 1.06E-05 |
| **8_125436151_G_A** | rs56056430 | [""""missense_variant:TRIB1""""] | 7.77E-05 |
| **8_125436158_G_T** |  | [""""missense_variant:TRIB1""""] | 1.06E-06 |
| **8_125436178_A_G** |  | [""""missense_variant:TRIB1""""] | 6.39E-06 |
| **8_125436178_A_T** |  | [""""missense_variant:TRIB1""""] | 1.06E-06 |
| **8_125436181_C_T** | rs555283941 | [""""missense_variant:TRIB1""""] | 1.06E-06 |
| **8_125436193_C_T** | rs775918973 | [""""stop_gained:TRIB1""""] | 1.06E-06 |
| **8_125436194_G_A** |  | [""""missense_variant:TRIB1""""] | 4.26E-06 |
| **8_125436206_A_G** |  | [""""missense_variant:TRIB1""""] | 1.06E-06 |
| **8_125436209_A_G** | rs200818822 | [""""missense_variant:TRIB1""""] | 2.13E-06 |
| **8_125436212_C_T** |  | [""""missense_variant:TRIB1""""] | 1.06E-06 |
| **8_125436226_C_A** |  | [""""missense_variant:TRIB1""""] | 2.13E-06 |
| **8_125436241_C_T** | rs200217434 | [""""missense_variant:TRIB1""""] | 8.51E-06 |
| **8_125436244_C_A** | rs55953723 | [""""missense_variant:TRIB1""""] | 4.26E-06 |
| **8_125436244_C_T** | rs55953723 | [""""missense_variant:TRIB1""""] | 4.26E-06 |
| **8_125436245_G_A** | rs151229265 | [""""missense_variant:TRIB1""""] | 6.49E-05 |
| **8_125436269_A_G** |  | [""""missense_variant:TRIB1""""] | 1.06E-06 |
| **8_125436270_C_A** | rs757119456 | [""""missense_variant:TRIB1""""] | 8.51E-06 |
| **8_125436275_C_T** | rs756032753 | [""""missense_variant:TRIB1""""] | 7.45E-06 |
| **8_125436278_C_T** | rs1162161939 | [""""missense_variant:TRIB1""""] | 2.13E-06 |
| **8_125436286_A_G** | rs774250787 | [""""missense_variant:TRIB1""""] | 6.39E-06 |
| **8_125436291_C_G** | rs1234672719 | [""""missense_variant:TRIB1""""] | 3.30E-05 |
| **8_125436292_C_T** |  | [""""missense_variant:TRIB1""""] | 3.19E-06 |
| **8_125436296_T_A** |  | [""""missense_variant:TRIB1""""] | 1.06E-06 |
| **8_125436298_C_T** | rs772430089 | [""""missense_variant:TRIB1""""] | 5.32E-06 |
| **8_125436299_G_A** | rs138022510 | [""""missense_variant:TRIB1""""] | 1.29E-04 |
| **8_125436302_G_A** | rs761181000 | [""""missense_variant:TRIB1""""] | 4.26E-06 |
| **8_125436313_C_T** | rs777006839 | [""""missense_variant:TRIB1""""] | 3.19E-06 |
| **8_125436314_G_A** | rs367629272 | [""""missense_variant:TRIB1""""] | 7.45E-06 |
| **8_125436320_C_T** | rs763855292 | [""""missense_variant:TRIB1""""] | 4.26E-06 |
| **8_125436325_G_A** | rs756273869 | [""""missense_variant:TRIB1""""] | 5.32E-06 |
| **8_125436325_G_T** | rs756273869 | [""""stop_gained:TRIB1""""] | 3.19E-06 |
| **8_125436335_C_G** |  | [""""missense_variant:TRIB1""""] | 4.26E-06 |
| **8_125436337_G_A** |  | [""""missense_variant:TRIB1""""] | 1.06E-06 |
| **8_125436341_C_A** |  | [""""missense_variant:TRIB1""""] | 1.06E-06 |
| **8_125436343_G_A** | rs755232835 | [""""missense_variant:TRIB1""""] | 3.19E-06 |
| **8_125436344_A_C** | rs1184690907 | [""""missense_variant:TRIB1""""] | 1.06E-05 |
| **8_125436348_C_G** |  | [""""missense_variant:TRIB1""""] | 1.06E-06 |
| **8_125436355_C_T** |  | [""""missense_variant:TRIB1""""] | 2.13E-06 |
| **8_125436358_C_T** | rs372024937 | [""""missense_variant:TRIB1""""] | 2.66E-05 |
| **8_125436359_C_G** | rs1030948982 | [""""missense_variant:TRIB1""""] | 2.13E-06 |
| **8_125436359_C_T** |  | [""""missense_variant:TRIB1""""] | 1.06E-06 |
| **8_125436364_T_G** |  | [""""missense_variant:TRIB1""""] | 2.13E-06 |
| **8_125436371_C_T** |  | [""""missense_variant:TRIB1""""] | 1.06E-06 |
| **8_125436385_G_A** | rs1025680568 | [""""missense_variant:TRIB1""""] | 6.39E-06 |
| **8_125436385_G_C** |  | [""""missense_variant:TRIB1""""] | 1.06E-06 |
| **8_125436415_G_A** |  | [""""missense_variant:TRIB1""""] | 2.13E-06 |
| **8_125436416_A_C** |  | [""""missense_variant:TRIB1""""] | 3.19E-06 |
| **8_125436418_C_T** |  | [""""stop_gained:TRIB1""""] | 8.51E-06 |
| **8_125436425_T_C** |  | [""""missense_variant:TRIB1""""] | 1.06E-06 |
| **8_125436431_A_C** | rs35454769 | [""""missense_variant:TRIB1""""] | 7.77E-05 |
| **8_125436436_C_G** | rs766477822 | [""""missense_variant:TRIB1""""] | 9.58E-06 |
| **8_125436436_C_T** | rs766477822 | [""""stop_gained:TRIB1""""] | 1.17E-05 |
| **8_125436448_G_C** |  | [""""missense_variant:TRIB1""""] | 3.19E-06 |
| **8_125436461_T_C** | rs992582240 | [""""missense_variant:TRIB1""""] | 5.32E-06 |
| **8_125436463_T_G** |  | [""""missense_variant:TRIB1""""] | 1.06E-06 |
| **8_125436464_T_A** |  | [""""missense_variant:TRIB1""""] | 1.06E-06 |

**Supplementary Table 6. PTV at *TM6SF2* with high impact or missense variants and CADD score ≥ 20.**

| **Variant ID** | **RSID** | **Consequence (Most severe by gene)** | **Population AF** |
| --- | --- | --- | --- |
| **19_19264740_C_T** | rs759316924 | [""""missense_variant:TM6SF2""""] | 2.34E-05 |
| **19_19264741_G_A** | rs1340544178 | [""""missense_variant:TM6SF2""""] | 4.26E-06 |
| **19_19264749_A_T** |  | [""""missense_variant:TM6SF2""""] | 3.19E-06 |
| **19_19264756_G_T** |  | [""""missense_variant:TM6SF2""""] | 1.06E-06 |
| **19_19264764_A_G** | rs1183815600 | [""""missense_variant:TM6SF2""""] | 2.02E-05 |
| **19_19264767_G_A** | rs757770572 | [""""missense_variant:TM6SF2""""] | 1.28E-05 |
| **19_19264768_C_A** | rs781763445 | [""""missense_variant:TM6SF2""""] | 2.13E-06 |
| **19_19264780_T_C** | rs768634850 | [""""missense_variant:TM6SF2""""] | 2.55E-05 |
| **19_19264791_A_C** | rs141184770 | [""""missense_variant:TM6SF2""""] | 7.20E-04 |
| **19_19264803_G_A** | rs905670454 | [""""missense_variant:TM6SF2""""] | 8.41E-05 |
| **19_19264813_G_A** |  | [""""missense_variant:TM6SF2""""] | 4.26E-06 |
| **19_19264818_C_T** | rs750920847 | [""""missense_variant:TM6SF2""""] | 2.02E-05 |
| **19_19264819_G_A** | rs756458112 | [""""missense_variant:TM6SF2""""] | 6.39E-06 |
| **19_19264821_T_G** |  | [""""missense_variant:TM6SF2""""] | 1.06E-06 |
| **19_19264833_G_A** |  | [""""missense_variant:TM6SF2""""] | 1.06E-06 |
| **19_19264836_C_T** | rs778992711 | [""""missense_variant:TM6SF2""""] | 8.51E-06 |
| **19_19264837_G_A** | rs1401840464 | [""""missense_variant:TM6SF2""""] | 1.17E-05 |
| **19_19264842_T_C** | rs1216044176 | [""""missense_variant:TM6SF2""""] | 1.06E-06 |
| **19_19264845_A_G** | rs748280759 | [""""missense_variant:TM6SF2""""] | 1.60E-05 |
| **19_19264851_G_A** |  | [""""missense_variant:TM6SF2""""] | 1.06E-05 |
| **19_19264850_AG_A** | rs1366147877 | [""""frameshift_variant:TM6SF2""""] | 3.19E-06 |
| **19_19264857_A_G** | rs1231996252 | [""""missense_variant:TM6SF2""""] | 1.06E-06 |
| **19_19264863_G_A** | rs745753117 | [""""missense_variant:TM6SF2""""] | 9.58E-06 |
| **19_19266494_C_T** | rs150912512 | [""""missense_variant:TM6SF2""""] | 1.44E-04 |
| **19_19266495_C_T** | rs199661804 | [""""missense_variant:TM6SF2""""] | 3.72E-05 |
| **19_19266500_C_A** |  | [""""missense_variant:TM6SF2""""] | 1.06E-06 |
| **19_19266512_A_C** |  | [""""missense_variant:TM6SF2""""] | 1.28E-05 |
| **19_19266515_A_G** | rs749156356 | [""""missense_variant:TM6SF2""""] | 1.28E-05 |
| **19_19266523_G_C** |  | [""""missense_variant:TM6SF2""""] | 2.13E-06 |
| **19_19266524_T_G** |  | [""""missense_variant:TM6SF2""""] | 2.13E-06 |
| **19_19266532_C_A** |  | [""""missense_variant:TM6SF2""""] | 1.06E-06 |
| **19_19266537_A_G** | rs1205611421 | [""""missense_variant:TM6SF2""""] | 5.32E-06 |
| **19_19266557_G_A** | rs374302893 | [""""missense_variant:TM6SF2""""] | 6.39E-06 |
| **19_19266581_G_A** |  | [""""missense_variant:TM6SF2""""] | 2.13E-06 |
| **19_19266595_C_T** |  | [""""missense_variant:TM6SF2""""] | 1.06E-06 |
| **19_19266596_A_G** | rs758547141 | [""""missense_variant:TM6SF2""""] | 2.13E-06 |
| **19_19266602_A_T** | rs757260859 | [""""missense_variant:TM6SF2""""] | 2.55E-05 |
| **19_19266610_C_T** |  | [""""splice_acceptor_variant:TM6SF2""""] | 3.19E-06 |
| **19_19267623_G_A** | rs755034726 | [""""stop_gained:TM6SF2""""] | 3.72E-05 |
| **19_19267637_G_A** | rs1257035007 | [""""missense_variant:TM6SF2""""] | 6.39E-06 |
| **19_19267638_C_A** | rs1456028329 | [""""missense_variant:TM6SF2""""] | 4.26E-06 |
| **19_19267647_C_T** | rs199926770 | [""""missense_variant:TM6SF2""""] | 2.15E-04 |
| **19_19267649_C_T** | rs370548010 | [""""missense_variant:TM6SF2""""] | 7.45E-06 |
| **19_19267649_C_G** |  | [""""missense_variant:TM6SF2""""] | 2.13E-06 |
| **19_19267650_G_A** | rs777516403 | [""""missense_variant:TM6SF2""""] | 7.45E-06 |
| **19_19267665_A_G** | rs770485209 | [""""missense_variant:TM6SF2""""] | 2.23E-05 |
| **19_19267689_C_CA** | rs1472997376 | [""""frameshift_variant:TM6SF2""""] | 5.32E-06 |
| **19_19267689_C_T** |  | [""""missense_variant:TM6SF2""""] | 7.45E-06 |
| **19_19267707_G_A** | rs1181949601 | [""""missense_variant:TM6SF2""""] | 1.06E-05 |
| **19_19267993_C_T** | rs766513047 | [""""missense_variant:TM6SF2""""] | 3.62E-05 |
| **19_19267993_C_A** | rs766513047 | [""""missense_variant:TM6SF2""""] | 1.28E-05 |
| **19_19267994_G_T** | rs181728218 | [""""synonymous_variant:TM6SF2""""] | 1.10E-04 |
| **19_19267994_G_A** | rs181728218 | [""""missense_variant:TM6SF2""""] | 1.70E-05 |
| **19_19267997_A_G** | rs1158032464 | [""""missense_variant:TM6SF2""""] | 3.19E-06 |
| **19_19268050_G_A** | rs186811910 | [""""missense_variant:TM6SF2""""] | 2.54E-04 |
| **19_19268053_C_T** | rs750038678 | [""""missense_variant:TM6SF2""""] | 5.00E-05 |
| **19_19268053_C_A** | rs750038678 | [""""missense_variant:TM6SF2""""] | 1.06E-06 |
| **19_19268054_G_A** | rs1216984470 | [""""missense_variant:TM6SF2""""] | 3.19E-06 |
| **19_19268690_G_C** |  | [""""missense_variant:TM6SF2""""] | 1.06E-06 |
| **19_19268709_G_T** | rs200148631 | [""""missense_variant:TM6SF2""""] | 3.45E-04 |
| **19_19268730_G_A** |  | [""""missense_variant:TM6SF2""""] | 1.06E-06 |
| **19_19268740_C_T** | rs58542926 | [""""missense_variant:TM6SF2""""] | 7.41E-02 |
| **19_19268742_GAGCTGTATTTGCCTTCCATGGTGCA_G** | rs746938988 | [""""splice_acceptor_variant:TM6SF2""""] | 4.26E-06 |
| **19_19268756_T_C** |  | [""""splice_acceptor_variant:TM6SF2""""] | 1.06E-06 |
| **19_19269704_A_G** | rs187429064 | [""""missense_variant:TM6SF2""""] | 1.30E-02 |
| **19_19269707_A_G** | rs202188864 | [""""missense_variant:TM6SF2""""] | 5.62E-04 |
| **19_19269716_A_T** |  | [""""missense_variant:TM6SF2""""] | 1.06E-06 |
| **19_19269716_A_G** | rs1412648831 | [""""missense_variant:TM6SF2""""] | 2.13E-06 |
| **19_19269734_C_G** | rs763650790 | [""""missense_variant:TM6SF2""""] | 6.39E-06 |
| **19_19269746_A_G** | rs1185700026 | [""""missense_variant:TM6SF2""""] | 1.70E-05 |
| **19_19269750_C_T** |  | [""""missense_variant:TM6SF2""""] | 1.06E-06 |
| **19_19269758_C_T** | rs369198322 | [""""missense_variant:TM6SF2""""] | 2.23E-05 |
| **19_19269759_G_A** | rs142056540 | [""""missense_variant:TM6SF2""""] | 1.06E-05 |
| **19_19269771_T_C** |  | [""""splice_acceptor_variant:TM6SF2""""] | 2.13E-06 |
| **19_19270186_C_T** | rs758530683 | [""""missense_variant:TM6SF2""""] | 4.04E-05 |
| **19_19270188_C_T** | rs969743528 | [""""missense_variant:TM6SF2""""] | 3.09E-05 |
| **19_19270191_G_A** | rs200162798 | [""""missense_variant:TM6SF2""""] | 6.49E-05 |
| **19_19270194_A_G** | rs754780228 | [""""missense_variant:TM6SF2""""] | 6.39E-06 |
| **19_19270195_T_C** |  | [""""missense_variant:TM6SF2""""] | 1.17E-05 |
| **19_19270203_T_C** |  | [""""missense_variant:TM6SF2""""] | 1.06E-06 |
| **19_19270204_A_G** |  | [""""missense_variant:TM6SF2""""] | 2.13E-06 |
| **19_19270210_G_A** |  | [""""missense_variant:TM6SF2""""] | 4.26E-06 |
| **19_19270211_G_T** | rs1455007681 | [""""stop_gained:TM6SF2""""] | 1.38E-05 |
| **19_19270215_T_A** | rs1206356828 | [""""missense_variant:TM6SF2""""] | 3.19E-06 |
| **19_19270225_C_T** |  | [""""missense_variant:TM6SF2""""] | 1.06E-06 |
| **19_19270227_T_A** | rs1400794525 | [""""missense_variant:TM6SF2""""] | 2.13E-06 |
| **19_19270228_C_G** |  | [""""missense_variant:TM6SF2""""] | 1.06E-06 |
| **19_19270228_C_T** |  | [""""missense_variant:TM6SF2""""] | 2.13E-06 |
| **19_19270245_A_G** |  | [""""missense_variant:TM6SF2""""] | 1.06E-06 |
| **19_19270248_C_G** | rs763226075 | [""""missense_variant:TM6SF2""""] | 9.58E-06 |
| **19_19270248_C_T** |  | [""""missense_variant:TM6SF2""""] | 2.13E-06 |
| **19_19270249_C_T** | rs771388055 | [""""missense_variant:TM6SF2""""] | 8.51E-06 |
| **19_19270249_C_A** | rs771388055 | [""""stop_gained:TM6SF2""""] | 4.26E-06 |
| **19_19270254_G_C** | rs765539044 | [""""missense_variant:TM6SF2""""] | 7.45E-06 |
| **19_19270254_G_A** | rs765539044 | [""""missense_variant:TM6SF2""""] | 5.32E-06 |
| **19_19270255_C_T** |  | [""""missense_variant:TM6SF2""""] | 1.06E-06 |
| **19_19270257_G_C** |  | [""""missense_variant:TM6SF2""""] | 1.06E-06 |
| **19_19270260_C_T** | rs201138158 | [""""missense_variant:TM6SF2""""] | 7.08E-04 |
| **19_19270261_G_A** | rs1167139822 | [""""missense_variant:TM6SF2""""] | 6.39E-06 |
| **19_19270273_C_T** | rs1395291293 | [""""missense_variant:TM6SF2""""] | 2.45E-05 |
| **19_19270273_C_G** |  | [""""missense_variant:TM6SF2""""] | 2.13E-06 |
| **19_19270347_C_G** |  | [""""missense_variant:TM6SF2""""] | 2.13E-06 |
| **19_19270389_C_T** |  | [""""missense_variant:TM6SF2""""] | 1.06E-06 |
| **19_19270400_A_C** |  | [""""missense_variant:TM6SF2""""] | 1.06E-06 |
| **19_19270406_A_G** |  | [""""missense_variant:TM6SF2""""] | 1.06E-06 |
| **19_19270407_G_A** | rs1205198625 | [""""missense_variant:TM6SF2""""] | 1.38E-05 |
| **19_19270415_A_G** |  | [""""missense_variant:TM6SF2""""] | 1.06E-06 |
| **19_19270418_G_A** | rs960855187 | [""""missense_variant:TM6SF2""""] | 2.13E-06 |
| **19_19270419_A_C** | rs749644443 | [""""missense_variant:TM6SF2""""] | 9.58E-06 |
| **19_19270440_A_G** | rs765124011 | [""""missense_variant:TM6SF2""""] | 2.13E-06 |
| **19_19271024_G_T** |  | [""""missense_variant:TM6SF2""""] | 1.06E-06 |
| **19_19271027_T_C** | rs527605147 | [""""missense_variant:TM6SF2""""] | 1.06E-05 |
| **19_19271031_G_A** |  | [""""missense_variant:TM6SF2""""] | 2.13E-06 |
| **19_19271049_C_T** | rs199996201 | [""""missense_variant:TM6SF2""""] | 1.81E-05 |
| **19_19271049_C_A** | rs199996201 | [""""stop_gained:TM6SF2""""] | 1.06E-06 |
| **19_19271060_A_C** |  | [""""missense_variant:TM6SF2""""] | 1.06E-06 |
| **19_19271066_T_C** | rs1469515809 | [""""missense_variant:TM6SF2""""] | 1.06E-06 |
| **19_19271116_C_T** | rs1272943322 | [""""stop_gained:TM6SF2""""] | 2.98E-05 |
| **19_19271120_A_G** |  | [""""missense_variant:TM6SF2""""] | 1.06E-06 |
| **19_19273119_A_C** |  | [""""splice_donor_variant:TM6SF2""""] | 1.06E-06 |
| **19_19273120_C_G** |  | [""""splice_donor_variant:TM6SF2""""] | 1.06E-06 |
| **19_19273122_G_T** |  | [""""missense_variant:TM6SF2""""] | 2.13E-06 |
| **19_19273124_G_A** | rs1188461427 | [""""missense_variant:TM6SF2""""] | 2.13E-06 |
| **19_19273127_A_G** |  | [""""missense_variant:TM6SF2""""] | 2.13E-06 |
| **19_19273133_G_A** | rs772045939 | [""""missense_variant:TM6SF2""""] | 4.26E-06 |
| **19_19273133_G_C** |  | [""""missense_variant:TM6SF2""""] | 3.19E-06 |
| **19_19273160_G_A** | rs576994101 | [""""missense_variant:TM6SF2""""] | 1.28E-05 |
| **19_19273160_G_C** | rs576994101 | [""""missense_variant:TM6SF2""""] | 2.13E-05 |
| **19_19273175_G_T** |  | [""""stop_gained:TM6SF2""""] | 4.26E-06 |
| **19_19273181_G_A** |  | [""""missense_variant:TM6SF2""""] | 1.06E-06 |
| **19_19273191_T_A** |  | [""""stop_gained:TM6SF2""""] | 3.19E-06 |
| **19_19273193_C_T** | rs867126720 | [""""missense_variant:TM6SF2""""] | 3.83E-05 |
| **19_19273203_G_C** | rs1209478584 | [""""missense_variant:TM6SF2""""] | 9.05E-05 |
| **19_19273203_G_A** |  | [""""missense_variant:TM6SF2""""] | 7.45E-06 |
| **19_19273203_G_T** | rs1209478584 | [""""missense_variant:TM6SF2""""] | 3.19E-06 |
| **19_19273211_T_C** | rs1460805562 | [""""missense_variant:TM6SF2""""] | 9.58E-06 |
| **19_19273215_T_C** | rs1417818061 | [""""start_lost:TM6SF2""""] | 1.06E-06 |
